# Comparative Medical Ecology of Gut Microbiomes in Major Neurodegenerative, Neurodevelopmental, and Psychiatric (NNP) Disorders

**DOI:** 10.1101/2025.03.16.25324039

**Authors:** Zhanshan (Sam) Ma, Yuting Qiao, Lianwei Li

## Abstract

This study provides a comprehensive medical ecology analysis of gut microbiome alterations in four neuropsychiatric disorders: Alzheimer’s disease (AD), Parkinson’s disease (PD), autism spectrum disorder (ASD), and the mood disorders (MDs) including major depressive disorder (MDD) (depression) and bipolar disorder (BD). Using diversity, heterogeneity, specificity, and AI/machine learning approaches, we examined microbiota and species-level dysbiosis by reanalyzing approximately 4000 gut microbiome samples from 23 independent studies on the four neuropsychiatric disorders. Key findings include: (i) Alpha-diversity increases across all disorders compared with healthy control, influenced by disease type and health status, while beta-diversity follows the Anna Karenina Principle (AKP), showing healthy individuals are more alike, while diseased individuals are not. Gamma and network diversity vary by disease type and species rarity-dominance spectrum. (ii) Unlike diversity, a common but limited approach ignoring species interactions, heterogeneity—considering both species abundance and interactions—remains understudied. TPLE (Taylor’s power law extension) ranked heterogeneity scaling as MD > ASD > PD ≈ H > AD, while TPLoN (TPL of network) showed an inverse order (H ≈ PD > ASD > MD > AD), except for AD. (iii) The SSD (species specificity and specificity diversity) framework not only identified disease-specific unique/enriched species (US/ES), which are promising candidates for probiotic development, but also identified key biomarker species crucial for the diagnosis of NNP disorders. In terms of species specificity, MD showed either negative correlations (with ASD, PD, and healthy controls) or insignificant correlation (with AD). In contrast, PD and AD were positively correlated but showed no correlation with ASD, potentially due to differences in patient age. These patterns align with intuitive perceptions of NNP disorders, though such perceptions have lacked microbiome-based evidence in existing literature. (iv) AI-Machine learning models achieved 80%-97% precision in diagnosing disorders with as few as top 25 most specific species. These results reveal shared and distinct microbiome features across neurodegenerative, neurodevelopmental, and psychiatric disorders, advancing understanding of their etiological mechanisms. The study highlights the potential of microbiome-based diagnostics and targeted therapies, such as probiotics and diagnostic biomarkers, to inform personalized interventions.

## Introduction

Neurodegenerative, neurodevelopmental, and psychiatric (NNP) disorders represent significant challenges in modern medicine, with complex etiologies involving genetic, environmental, and microbial factors. Among these, the gut microbiome has emerged as a critical modulator of neurological and psychiatric health through the microbiota-gut-brain axis. This bidirectional network links the central nervous system (CNS) with gut microbial communities, influencing neural development, immune function, and metabolic processes. Disruptions in this axis have been implicated in various NNP disorders, highlighting the gut microbiome as a potential diagnostic marker and therapeutic target. Here, we provide a comprehensive analysis of the gut microbiome’s role in five prevalent NNP disorders—Autism Spectrum Disorder (ASD), Major Depressive Disorder (MDD), Bipolar Disorder (BD), Alzheimer’s Disease (AD), and Parkinson’s Disease (PD)—and explore how microbial dysbiosis contributes to their pathophysiology.

Autism Spectrum Disorder (ASD), a neurodevelopmental condition affecting approximately 2% (1 in 54) children worldwide, is characterized by social communication deficits, restricted interests, and repetitive behaviors (Lord et al., 2018). While genetic factors contribute to its pathogenesis, environmental influences, particularly the gut microbiome, are increasingly recognized as modulators of ASD development. Individuals with ASD often exhibit reduced microbial diversity and an overrepresentation of pathobionts, such as *Clostridium* species (Sharon et al., 2019). This dysbiosis exacerbates gastrointestinal symptoms, which may influence behavioral severity (Korteniemi et al., 2023). Microbial imbalance in ASD contributes to neuroinflammation and disrupted brain function. For example, lipopolysaccharides (LPS) produced by gut bacteria can translocate into the bloodstream, triggering systemic inflammation (Wan et al., 2022). Reduced microbial production of short-chain fatty acids (SCFAs), particularly butyrate, compromises gut epithelial integrity and promotes neuroinflammatory responses (Lord et al., 2018). Animal studies have shown that transplanting ASD-associated microbiota into germ-free mice induces autism-like behaviors, supporting a causal role for the microbiome (Sharon et al., 2019). These findings have spurred interest in microbiome-targeted interventions, such as probiotics and dietary modifications, to alleviate ASD symptoms (Hirota & King, 2023).

Major Depressive Disorder (MDD), a leading cause of disability worldwide, is characterized by persistent sadness, cognitive and somatic symptoms, and disrupted sleep patterns (Smith, 2014). Recent evidence implicates the gut microbiome in MDD pathogenesis. Individuals with MDD exhibit reduced microbial diversity, with a depletion of beneficial microbes like *Faecalibacterium prausnitzii* and an overgrowth of pro-inflammatory species (Simpson et al., 2021). Altered SCFA levels disrupt the blood-brain barrier, increasing neuroinflammation and depressive symptoms (Liu et al., 2023). The overlap between MDD and gastrointestinal disturbances, such as irritable bowel syndrome (IBS), further supports the gut-brain connection (Aziz *et al*. 2021). Microbiome-targeted therapies, including probiotics and prebiotics, have shown promise in alleviating depressive symptoms by modulating gut-brain signaling pathways. For instance, *Lactobacillus* and *Bifidobacterium* supplementation enhances serotonergic activity and reduces inflammation (Anderson et al., 2024). However, further research is needed to optimize these interventions (Alli et al., 2022).

Bipolar Disorder (BD), marked by extreme mood swings, involves genetic, environmental, and neurochemical dysregulation (Harrison et al., 2018). Gut dysbiosis in BD patients, characterized by reduced *Faecalibacterium* and increased *Escherichia-Shigella*, exacerbates systemic inflammation and neurochemical imbalances (Li et al., 2022). The microbiota-gut-brain axis influences mood regulation through neurotransmitter modulation, such as serotonin and dopamine metabolism (Sublette et al., 2021). Deficits in SCFA production and elevated LPS levels in BD contribute to neuroinflammation and oxidative stress (McGuinness et al., 2022). Probiotic supplementation has shown potential in alleviating depressive symptoms, though its effects on manic episodes remain understudied (Tondo et al., 2017). Further research is needed to explore microbiome-based therapies for mood stabilization and inflammation reduction (Sublette et al., 2021).

Alzheimer’s Disease (AD), the most common neurodegenerative disorder, is characterized by cognitive decline, memory loss, and amyloid-beta and tau pathology (Scheltens et al., 2021). The gut microbiome plays a pivotal role in AD progression, with patients exhibiting increased pro-inflammatory taxa like *Escherichia* and *Proteobacteria* (Chen et al., 2022). Dysbiosis contributes to neuroinflammation and amyloid-beta deposition, accelerating neurodegeneration (Graff-Radford et al., 2021). Reduced SCFA levels in AD disrupt the blood-brain barrier, facilitating neurotoxic substance entry and microglial activation (Zhang et al., 2023). Microbial-derived metabolites, such as LPS, exacerbate cognitive decline (Ferreiro et al., 2023). Preclinical studies suggest that microbiome-targeted therapies, including fecal microbiota transplantation (FMT) and dietary interventions, may mitigate cognitive deficits and amyloid burden (Ferreiro et al., 2023). However, clinical trials are needed to assess their safety and efficacy (Zhang et al., 2023).

Parkinson’s Disease (PD), a progressive neurodegenerative disorder, is characterized by motor symptoms and dopaminergic neuron loss in the substantia nigra (Moussa et al., 2017). The gut microbiome’s role in PD is increasingly recognized, with patients exhibiting reduced *Prevotella* and increased *Enterobacteriaceae* (Sampson et al., 2020). These shifts influence the gut-brain axis, promoting neuroinflammation and alpha-synuclein aggregation, a key PD hallmark (López et al., 2022). Gastrointestinal symptoms, such as constipation, often precede motor symptoms, suggesting early microbial involvement in PD pathogenesis (Sampson et al., 2020). SCFAs, particularly butyrate, play a protective role in maintaining gut barrier function and modulating neuroinflammation (Gao et al., 2021). Microbiome-targeted therapies remain a promising area of investigation for PD management.

The growing body of evidence linking the gut microbiome to NNP disorders highlights the potential of the microbiota-gut-brain axis as a therapeutic target. While each disorder—ASD, MDD, BD, AD, and PD—presents unique microbial signatures, common features such as reduced SCFA production and increased pro-inflammatory taxa suggest shared pathogenic mechanisms. Advances in microbiome research, coupled with multi-omics approaches integrating genomic, metagenomics, metabolomic, and neuroimaging data, are paving the way for a more comprehensive understanding of these complex disorders. This study is aimed to conduct comparative medical ecology analyses of the gut microbiomes associated with major NNP disorders. Understanding the differences and commonalties in gut microbiome composition among various neurodegenerative, neurodevelopmental, and psychiatric disorders is crucial for several reasons. First, these microbial signatures offer valuable insights into the underlying pathophysiology of each disease. For instance, while the dysbiosis in AD and PD is characterized by increased pro-inflammatory bacteria, such as *Proteobacteria* and *Firmicutes*, ASD may involve distinct microbial alterations, such as an overrepresentation of *Clostridium* species (Sharon et al., 2019; Liu et al., 2023). Identifying these disease-specific microbiota profiles allows for more precise diagnostic biomarkers, which could lead to earlier detection and better prognostic assessment. Second, comparing the gut microbiome of disease-affected individuals with that of healthy controls can highlight microbial pathways that are either disrupted or protective in different neurological conditions. For example, a reduction in beneficial bacteria like *Bifidobacterium* and *Lactobacillus* and a concomitant increase in pathobionts such as *Escherichia-Shigella* have been observed in both Major Depressive Disorder (MDD) and Bipolar Disorder (BD) (Simpson et al., 2021; Tondo et al., 2017). These findings suggest that while the gut microbiome is disrupted in both disorders, the specific microbial shifts might contribute to different disease outcomes, from depressive episodes in MDD to mood instability in BD. Such insights help tailor more targeted interventions, such as probiotics or dietary modifications, based on the individual’s unique microbial profile (McGuinness et al., 2022). Moreover, understanding these differences in gut microbiome composition has profound implications for the development of microbiome-based therapies. Given the shared features across disorders, including neuroinflammation and altered SCFA production, therapies targeting these microbial signatures could provide a generalized approach to treating multiple disorders. However, the disease-specific nuances in microbiota composition underscore the need for personalized treatment strategies that take into account not only the disease in question but also the patient’s microbiome. Finally, the comparative analysis of gut microbiome differences between disease and healthy states can provide insights into the broader role of microbial ecosystems in maintaining brain health.

While dysbiosis is a hallmark of many neurodegenerative and psychiatric conditions, a healthy, balanced microbiome appears to protect against neuroinflammation, modulate immune responses, and support cognitive and emotional well-being (Liu et al., 2023; Anderson et al., 2024). As we gain a deeper understanding of how specific microbial populations influence the central nervous system, it is increasingly clear that preserving or restoring gut microbial health may not only help in managing the symptoms of these diseases but may also serve as a preventive strategy for individuals at risk. In conclusion, advancing our knowledge of gut microbiome differences across these NNP disorders, as well as the disparities between disease states and health, is pivotal for the development of innovative, microbiome-targeted therapies. As research evolves, microbiome-based interventions hold the potential to complement existing treatments, offering more effective, personalized therapeutic options for these complex, multifactorial conditions.

Dysbiosis, also a key feature of ecosystem dynamics, fundamentally represents a loss of balance or disequilibrium within ecosystems. As such, medical ecology analysis holds particular significance for comparing NNP disorders. The medical ecology of human microbiome-associated diseases is an interdisciplinary field that integrates medical microbiology, clinical medicine, computational biology, metagenomics bioinformatics, and theoretical ecology—particularly community ecology (Ma & Zhang 2022). We posit that the first principles underlying microbiome-associated diseases are inherently ecological in nature. In this study, we focus on applying ecological theories and methods to conduct a comprehensive comparison of NNP disorders. These include analyses of diversity (incorporating network diversity), heterogeneity (network heterogeneity), dominance (dominance network), specificity (leveraging the composition heterogeneity to detect treatment-specific unique/enriched species), and AI-machine learning approaches. By leveraging these ecological frameworks, we aim to uncover deeper insights into the microbiome’s role in NNP disorders. We accomplished our objectives by reanalyzing extensive datasets of NNP disorders, encompassing approximately 4,000 gut microbiome samples from both patients and healthy controls.

## Material and Methods

### Data Collection and Bioinformatics Analysis

This study analyzed 23 publicly available 16S rRNA sequencing datasets from the NCBI Sequence Read Archive (SRA) to investigate the gut microbiome across five neurological disorders: Autism Spectrum Disorder (ASD), Depression (DEP), Bipolar Disorder (BD), Alzheimer’s Disease (AD), and Parkinson’s Disease (PD). These datasets, representing diverse global populations, provided a comprehensive comparison of microbial profiles between affected individuals and healthy controls. A detailed summary of the datasets, including Dataset IDs, sample sizes, geographical origins, and publication years, is provided in Table S1. Datasets were selected based on the following criteria: (i) inclusion of both disease and healthy control groups, (ii) availability of raw 16S rRNA sequencing data in the NCBI SRA database, and (iii) clear labeling of the neurological disorder represented. Of the 23 datasets, 21 were supported by published articles describing their research context, methodologies, and findings. Two datasets (PRJNA596190 and PRJNA776170) lacked associated publications but were included due to sufficient metadata and sequencing information. For analytical purposes, datasets related to Depression (DEP) and Bipolar Disorder (BD) were grouped under the Mood Disorders (MDs) category, enabling the examination of shared gut microbiome features reflective of their overlapping clinical and microbial characteristics.

Raw sequencing data were downloaded from the NCBI SRA using the SRA Toolkit (version 3.0.7), with the prefetch and fastq-dump commands employed to retrieve FASTQ files. The data were processed using a bioinformatics pipeline comprising Trimmomatic (v0.39), Kraken2 (v2.1.2), and Bracken (v2.6), with the 16S Greengenes database (March 25, 2020) as the reference. Quality control steps included the removal of sequencing artifacts and low-quality reads. Trimmomatic (v0.39) was used to trim adapter sequences and low-quality regions from the 5’ and 3’ ends of reads, retaining only high-quality segments. Reads with a Phred quality score below 30 or shorter than 50 base pairs after trimming were discarded.

Taxonomic classification and Operational Taxonomic Unit (OTU) identification were performed using Kraken2 (v2.1.2), a *k*-mer-based tool that assigns taxonomy to sequence reads by comparing them to a reference database. Kraken2’s output was refined using Bracken (v2.6), which employs a probabilistic model to estimate the relative abundance of taxa at the genus and species levels, adjusting for biases in read length and classification uncertainty. This step enhanced the accuracy of taxonomic abundance estimates, particularly for datasets with varying sequencing depths. Following Kraken2 and Bracken processing, OTUs were generated by aggregating sequences at the genus or species level, depending on the analysis requirements. The resulting OTU tables, containing taxonomic assignments and their corresponding abundances, were formatted for downstream analysis using KrakenTools and R-software (v4.1.2). A summary of the OTU tables and dataset information is provided in Table S1. Furthermore, Figure S1 demonstrates the organization of the 23 datasets into NNP treatments (including AD, ASD, MD, and PD) and their corresponding healthy controls for each medical ecology analysis outlined in Figure 1 below. This figure highlights the medical ecology methods employed in this study.

**Fig 1.**
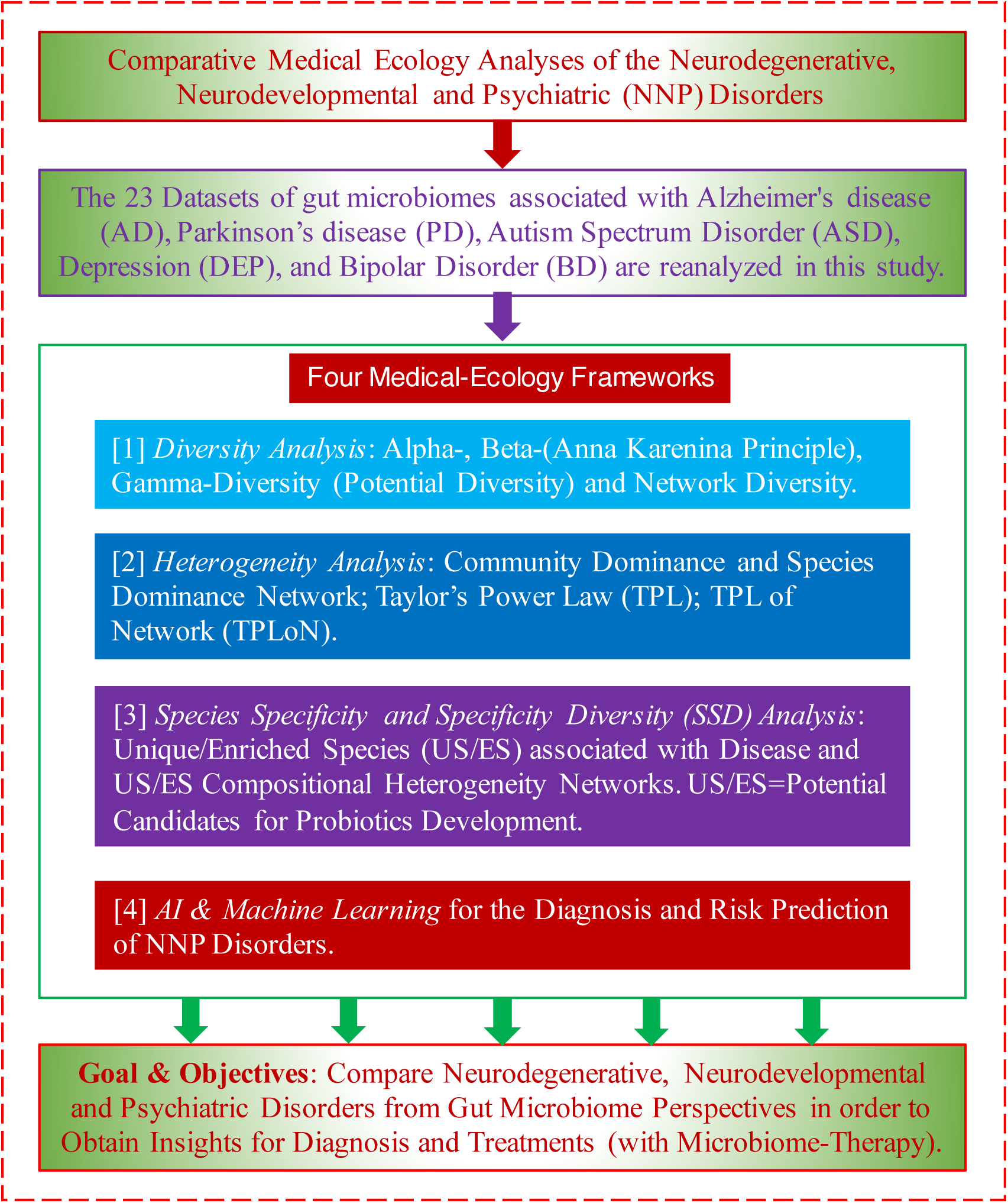
The study design for comparing the gut microbiomes associated with four NNP disorders

### Comparative Analysis of Alpha-, Beta-, and Gamma-Diversity

#### Alpha-Diversity with Hill Numbers and Non-parametric Two-Way ANOVA

Alpha-diversity, often referred to simply as diversity or biodiversity, is a de facto standard analysis in virtually all microbiome studies. In this study, the bacterial diversity of gut microbiomes is assessed and compared across four neurodegenerative, neurodevelopmental, and psychiatric (NNP) disorders —Alzheimer’s Disease (AD), Autism Spectrum Disorder (ASD), Mood Disorders (MD), and Parkinson’s Disease (PD)—as well as a healthy control group. The analysis employs Hill numbers at diversity orders *q* = 0-2. While numerous diversity measures (metrics or indices) exist in the literature, there is now a general consensus that Hill numbers provide a unifying framework for nearly all of them, offering the most appropriate metrics for assessing alpha-diversity (Ellison 2010; Chao et al. 2012, 2014). Specifically, at *q* = 0, the Hill number corresponds to species richness; at *q* = 1, it represents the effective number of commonly abundant species; and at *q* = 2, it reflects the effective number of dominantly abundant species.

To compare the alpha-diversity of gut microbiomes across NNP disorders, we employed a non-parametric two-way ANOVA using the Scheirer-Ray-Hare test, followed by Tukey’s Honest Significant Difference (TukeyHSD) post hoc tests for pairwise comparisons. The TukeyHSD test was applied to account for multiple comparisons, with statistical significance determined using a threshold of *P*-value < 0.05. Additionally, corrections for false discovery rates (FDR) were implemented to ensure robust and reliable results.

Hill numbers, also known as the effective number of species or true diversity indices, are defined for a given order *q* as follows:

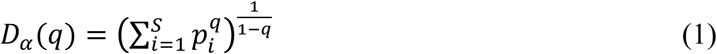

where 𝑝_𝑖_ represents the relative abundance of species *i*, and *S* is the total number of species.

To evaluate the effects of Disease Type (AD, ASD, MD, PD) and Health Status (healthy *vs*. diseased) on alpha-diversity, a non-parametric two-way ANOVA is performed using the Scheirer-Ray-Hare test. This test is particularly suitable for analyzing non-normally distributed data, which is often encountered in microbiome studies. The statistical model for this test is as follows:

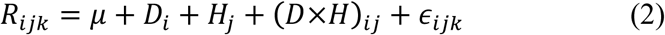

where *R*_ij_ represents the ranks of the alpha-diversity values for the *i-*th Disease Type and *j*-th Health Status, 𝜇 is the overall mean rank, 𝐷_𝑖_ is the effect of the *i*-th Disease Type (AD, ASD, MD, PD), 𝐻_j_ is the effect of *j*-th Health Status (𝐷×𝐻)_ij_ is the interaction term between Disease Type and Health Status, є_𝑖jk_ is the residual error term.

The Scheirer-Ray-Hare test extends the Kruskal-Wallis test to accommodate two-way factorial designs, making it ideal for assessing the main effects of Disease Type and Health Status, as well as their potential interaction. After performing the test, Tukey’s Honest Significant Difference (TukeyHSD) post-hoc tests are applied for pairwise comparisons to identify specific differences between groups.

*Post-hoc* multiple comparisons were conducted using the aov function in R, followed by Tukey’s Honest Significant Difference (TukeyHSD) test, which adjusts for multiple comparisons and calculates 95% family-wise confidence intervals for each pairwise comparison. The TukeyHSD test is based on the following formula:

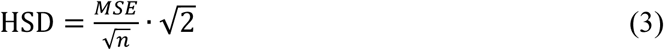

where *MSE* is the mean square error from the ANOVA table, and *n* is the sample size for each group. Significance is determined using a threshold of P-value < 0.05, with corrections for false discovery rates (FDR) to control for multiple comparisons. This approach ensures robust and reliable identification of significant differences in alpha-diversity across the studied groups.

#### Beta-Diversity for Testing the Anna Karenina Principle (AKP) Effects

The Anna Karenina Principle (AKP) posits that healthy microbiomes tend to be relatively stable and similar across individuals, whereas diseased microbiomes exhibit greater variability and instability. Inspired by Tolstoy’s observation that “all happy families are alike; each unhappy family is unhappy in its own way,” this principle suggests that dysbiosis (microbial imbalance) manifests uniquely in each diseased individual (Zaneveld et al. 2017; Ma 2020). In microbiome-associated diseases, compromised host immunity or environmental stressors can disrupt microbiome stability, leading to increased heterogeneity in community composition. Beta-diversity metrics, particularly when framed within the context of Hill numbers, provide a quantitative measure of this variability. Higher beta-diversity in diseased groups than in healthy controls serves as a hallmark of AKP effects, while lower beta-diversity indicates anti-AKP dynamics. Statistically insignificant differences in beta-diversity between healthy and diseased groups suggest non-AKP effects (Ma 2020). This framework allows for a nuanced understanding of microbiome variability in health and disease states.

To investigate the Anna Karenina Principle (AKP) in NNP disorders, we utilized Hill numbers as a robust and unifying framework for quantifying alpha-, beta-, and gamma-diversity. Specifically, we assessed AKP effects by examining beta-diversity within the framework of Hill numbers.

Gamma-diversity, which represents the total diversity of pooled communities, is defined as:

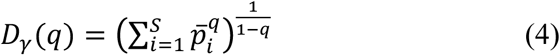

where 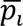 is the relative abundance of species *i* in the pooled assemblage of *N* communities. Beta-diversity, which measures the heterogeneity between communities, was multiplicatively partitioned as:

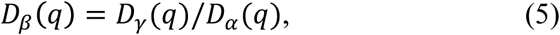

where 𝐷_𝛼_(𝑞) is the average alpha-diversity across *N* communities, as defined in Eqn. (1).

This multiplicative partitioning aligns with the recent consensus on the advantages of Hill numbers for biodiversity analysis, as multiplicative partitioning of beta-diversity is considered more appropriate than additive partitioning (Chao et al. 2012, 2014).

To detect AKP effects, we compared beta-diversity between healthy (H) and diseased (D) groups. Higher beta-diversity in the D group indicates increased heterogeneity in community composition, a hallmark of AKP. Statistical significance was assessed using a non-parametric Wilcoxon test. If pairwise beta-diversity within the H group was significantly lower than in the D group (*P*-value < 0.05), AKP effects were inferred. Conversely, anti-AKP effects (the opposite trend) were inferred if beta-diversity in the H group was significantly higher, and non-AKP effects were inferred when no significant difference was detected.

#### Gamma-diversity—Diversity-scaling analysis and potential (dark) diversity

Ma (2018) extended the classic species-area relationship (SAR) using Hill numbers, transforming it into the diversity-area relationship (DAR). The SAR, first discovered in the 19th century (Watson 1835), predicts that the number of species (S) in a region scales logarithmically with the area size (A). The DAR generalizes this relationship by replacing species richness (S) with diversity (D) measured in Hill numbers, expressed as:

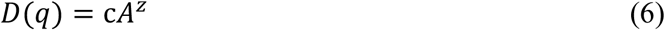

where *D*(*q*) is the diversity in Hill numbers of order *q*, *A* is the area size, and *c* and *z* are DAR parameters. Here, *c* approximates the average alpha diversity across communities, while *z* represents the rate of diversity change across regions. In other words, while *z* can be considered a proxy for beta-diversity and *c* a proxy for alpha-diversity, *D(q)* essentially serves as a proxy for gamma diversity.

Previous attempts to extend SAR with general diversity measures were less successful because the metrics used lacked the superior properties of Hill numbers. Hill numbers provide a unified framework for biodiversity measurement, akin to a conversion mechanism (similar to the gold-to-USD exchange in the Bretton Woods system), enabling the expression of diversity as effective numbers of species or species equivalents—a feature absent in other diversity metrics.

Ma (2018, 2019) introduced the concept of potential (dark) diversity, which encompasses both species present in local communities and those absent locally but present in the regional species pool, with the potential to colonize local communities through regional migration. This potential diversity represents the total species richness at regional or global scales, such as the total microbial species in Parkinson’s disease (PD) patients. Traditional diversity analyses often miss this information due to the extensive heterogeneity of human microbiomes and the potential for microbial transfer across human populations on ecological and evolutionary timescales, making it infeasible to simply sum species from all individuals. To estimate potential diversity, Ma (2018) extended the diversity-area relationship (DAR) power function (Eqn. 6) with a power-law with exponential cutoff (PLEC):

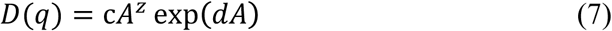

where *d* is a third parameter, typically negative, acting as a taper-off parameter that constrains exponential decay [exp(*d*A)]. This decay eventually overwhelms the power-law behavior (unlimited growth) at very large area sizes (A). From Eqn. (7), Ma (2018) derived the equation for computing the potential diversity, *D*_max_(*q*), as:

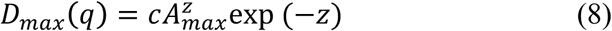

where 𝐴_𝑚𝑎𝑥_ = (−𝑧/𝑑) is the area size when diversity accrual reaches its maximum regionally or globally.

#### Network Diversity

Ohlmann et al. (2019) propose a comprehensive approach to measure network diversity at local (α-diversity), regional (γ—diversity), and between-sample (β—diversity) levels, accounting for the probabilistic nature of biotic interactions and species abundances. Their framework highlights the importance of analyzing network diversity across multiple levels of species aggregation, from individual species to broader trophic groups, to better understand network structure and dynamics. Ma & Li (2024) apply Ohlmann et al. (2019) framework to microbiome networks, specifically animal gastrointestinal microbiomes. Ma & Li (2024) discussed the significance of Hill numbers as a unifying system for biodiversity metrics, drawing parallels between ecological and economic systems. They note that Hill numbers, based on Renyi entropy and rooted in economic concepts like the “numbers equivalent,” provide a standardized framework akin to linking currencies to a gold standard.

The network diversity (ND) metrics provide a framework for quantifying biodiversity in ecological networks by integrating species interactions and their structural dynamics. These metrics can be categorized into three sub-metrics: (i) ND of species relative abundances (NDSA), (ii) ND of link probability (NDLP), and (iii) ND of link abundances (NDLA). NDSA focuses on the diversity of species within a network, weighted by their relative abundances. NDLP quantifies the diversity of potential interactions (links) between species, considering the probabilistic nature of these interactions. This sub-metric captures the variability in interaction likelihoods, reflecting the complexity and uncertainty inherent in ecological networks. NDLA measures the diversity of interaction strengths, incorporating the weight or frequency of links between species. This metric highlights the importance of strong versus weak interactions in shaping network structure and dynamics. Together, these sub-metrics provide a comprehensive toolkit for analyzing ecological networks at multiple levels, from species abundances to interaction probabilities and strengths. By unifying these dimensions, network diversity metrics enable ecologists to study biodiversity patterns, compare networks across environments, and understand how species interactions respond to environmental gradients and temporal changes. The network diversity framework proposed in Ohlmann et al. (2019) and demonstratively applied in Ma & Li (2024), marks a paradigm shift in biodiversity research, emphasizing the critical role of species interactions in ecosystem functioning and resilience. To some extent, network diversity metrics can serve as a bridge between diversity and heterogeneity studies. The mathematical formulations for the network diversity (ND) developed by Ohlmann et al. (2019) are also based on Hill numbers [Eqn. (1), (4), (5)]. For detailed computational procedures and programs for implementing the NDSA, NDLP, and NDLA, refer to Ma & Li (2024).

### Heterogeneity analysis

Heterogeneity and diversity are frequently conflated concepts. As Aaron Ellison aptly puts it, “a zoo is diverse, while an ecosystem is heterogeneous.” When a zookeeper assesses diversity, they simply count the different kinds of animals, without expecting interactions between them due to barriers like fences. On the other hand, measuring heterogeneity should account for both the interactions among species and the relationships between species and their heterogeneous habitats.

While measuring diversity using Hill numbers is a relatively well-established practice in community ecology, quantifying heterogeneity remains a significant challenge. Measuring diversity typically relies on entropy, whereas variance is often used to assess heterogeneity. For instance, one of the simplest heterogeneity metrics is the variance-to-mean ratio. However, using variance as a measure of heterogeneity is far from ideal, as variance primarily serves as a proxy for the degree of data dispersion. At best, it can only indirectly reflect the effects of species interactions, rather than providing a direct measure of those interactions *per se*.

In this study, we apply three methods for measuring heterogeneity: TPLE (Taylor’s power law extensions) (Ma 2015, Taylor 2019), TPLoN (TPL of network) (Ma 2025) and dominance analysis (Ma & Ellison 2019, 2024).

#### Dominance analysis

Ma & Ellison (2018) introduce a unified framework for measuring community dominance and species dominance in ecological systems, particularly for investigating dominance-stability relationships. The framework builds on Lloyd’s (1967) mean crowding index to quantify dominance at both the community and species levels, offering an alternative approach to classic diversity-stability relationships.

Community dominance (*D*_c_) measures the overall unevenness of species distributions within a community. It is calculated as a function of mean population abundance and variance, reflecting how dominance structures the community. A high *D_c_* indicates a community dominated by a few species, while a low *D*_c_ suggests a more even distribution. This metric is closely related to traditional diversity indices like Gini-index and Simpson’s diversity index but provides a more nuanced understanding of dominance dynamics.

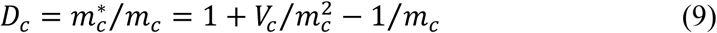

where s_C_ is mean community crowding and is derived as:

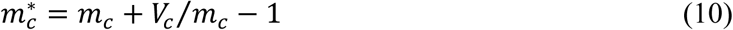

*m*_c_ is the mean of the population abundances (sizes) across species (*i.e*., per species) in the community, and *V*_c_ is the corresponding variance. There is a rigorous linear function relationship between community dominance (*D*_c_) and Simpson’s diversity index, which is a function of familiar Gini-index in economics. Hence, *D*_c_ is well-correlated with other measures of diversity. One advantage of *D*_c_ is that it can be applied to define a dominance index for each species in the community.

Species dominance (*D*_s_) focuses on the relative contribution of individual species to community structure. It quantifies how much a species dominates compared to the community as a whole, using the difference between community dominance and a species-specific dominance distance. As mentioned previously, community dominance is a function of the familiar Gini-index in economics, while species dominance is analogous to assigning a Gini-index to each individual within a country, whereas the Gini-index itself is only applicable at the country level. The species dominance is defined with the following equation (Ma & Ellison 2018):

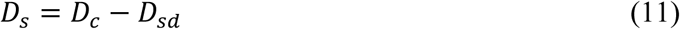

where *D_sd_* is the *species dominance distance* and represents the distance between the “gravity center” of community dominance and the focal species positon in an imaginary sphere of community. *D_sd_* is defined, similar with community dominance formula (Eqn. 14):

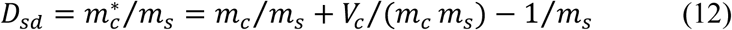

where *m_s_* is the population abundance (size) of the focal species of interest (*s*) in the community. Obviously, *D_s_* is species-specific, and it represents the dominance of a specific (focal) species in the community: more dominant species corresponds to greater values of *D_s_*.

This approach allows for the identification of key species, such as the most dominant OTU (MDO) or most abundant OTU (MAO), which play critical roles in shaping community stability and function. By performing permutation tests for species dominance, one can identify species that are significantly enriched or depleted in diseased (*e.g*., Parkinson’s disease) or healthy conditions.

Species dominance can also be utilized to build species dominance networks (SDNs), similar to constructing species correlation networks using species abundances. By analyzing SDNs, researchers can identify core-periphery structures, trio motifs, and high-salience skeletons, revealing how dominant species influence community stability. Indeed, SDNs serve as the primary tool for investigating the dominance-stability relationship, offering advantages over the classic diversity-stability relationship paradigm.

In this study, we apply dominance analysis for two purposes. First, we conduct community dominance analysis with non-parametric two-way ANOVA, similar with the previous alpha-diversity analysis. Second, we construct species co-occurrence networks (SCNs) by substituting traditionally used species abundances with species dominance. Unlike species abundances, which are compositional data, species dominance is not subject to compositional effects. Consequently, correlation coefficients derived from species dominance are free from the biases that can distort the identification of negative correlations (Ma & Li, 2024). By utilizing species dominance, we can apply computationally efficient Spearman’s correlation coefficients to build SCNs in this study.

#### Taylor’s power law and its extensions

The Taylor Power Law (TPL) was first discovered by L. R. Taylor (1961) during his study of insect populations, particularly their spatial distribution (aggregation). TPL describes the relationship between the mean population abundance (*m*) and its corresponding variance (*V*) using a power function, expressed as:

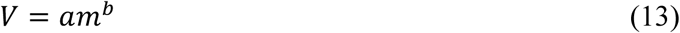

where *a* and *b* are TPL parameters. The parameter *b* is regarded as the heterogeneity scaling parameter, as it represents the slope of the variance-mean (*V-M*) relationship when plotted on a logarithmic scale, *i.e.,*

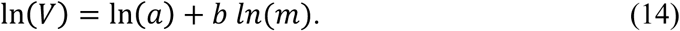

When *b*>1, population spatial distribution is aggregated (heterogeneous), *b*=1, random, and *b*<1 uniform or regular. Ma (1991) further proposed the concept of population aggregation critical density (PACD) or *m*_0_,

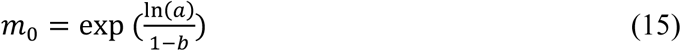

where *a* and *b* are TPL parameters. The significance of *m_0_* lies in its role as an estimator of a critical threshold where the spatial distribution transitions to randomness. This threshold marks a tipping point between aggregated and uniform distributions, akin to phase transitions observed in physical systems.

Ma (2015) extended the Taylor Power Law (TPL) to the community level by introducing four TPL extensions (TPLEs): Type-I TPLE for measuring community spatial heterogeneity, Type-II TPLE for assessing community temporal stability, Type-III TPLE for evaluating mixed-species population spatial aggregation, and Type-IV TPLE for analyzing mixed-species population temporal stability. While these TPLEs retain the same power function form as the original TPL, they differ in how the mean (*m*), variance (*V*), and corresponding TPLE parameters are quantified and interpreted. Furthermore, the TPLE parameters can be utilized to calculate the community heterogeneity threshold (CHT), which serves as the community-level counterpart to the population aggregation critical density (PACD) observed at the population scale. This threshold provides a critical point for understanding shifts in community spatial and temporal dynamics, offering insights into the transition between different states of community heterogeneity and stability.

As previously discussed, using variance or variance-mean modeling to measure heterogeneity is far from ideal, as it does not directly account for species interactions. To address this limitation, Ma (2025) extended Taylor’s Power Law (TPL) to complex ecological networks. The key distinctions between TPL (or TPLE) and TPLoN (Taylor’s Power Law of Networks) are as follows: (i) A species co-occurrence network is first constructed for groups of microbiota, such as comparing microbiome samples from Parkinson’s disease (PD) patients with those from healthy controls. Consequently, the scale for TPLoN operates at the meta-community or regional level. (ii) The concept of weighted species connectedness (WC) within the species co-occurrence network is introduced and quantified. WSC serves as a direct measure of species interactions, which are central to understanding heterogeneity. (iii) TPL is applied to the mean (*m*) and variance (*V*) of WC, providing a novel framework for analyzing ecological heterogeneity in networked systems. The implementation of TPLoN (Taylor’s Power Law of Network) can be decomposed as the following five steps:

*Step 1*: Compute Microbial OTU Correlations for building species co-occurrence networks (SCN) with algorithms such as Spearman’s correlation coefficients or SparCC algorithm. Perform FDR (false discovery rate) control to filter out potentially false OTU correlations.

*Step 2*: Define Species Connectedness (*C*_i_)

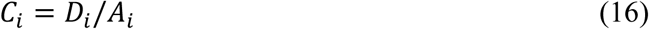

where *D*_i_ = network degree of species (*i*) or the number of significant correlations or links.

*A*_i_= mean abundance of species (*i*) across all samples. Connectedness represents the number of significant correlations per individual of species (*i*), reflecting its role in the network.

*Step 3*: Define Weighted Species Connectedness (WC) of species *i*,

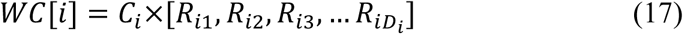

where species connectedness *C_i_* is defined in the previous step, *R_ij_* represents for the correlation coefficient between species *i* and*j, j*=1, 2, … *D_i_,* and *D_i_* is the degree of species *i,* or the number of nearest neighbors (*i.e*., direct links) species *i* is linked to. In other words, the *weighted connectedness* of species *i, WC*[*i*], is a *vector* of the product of the species connectedness (*C_i_*) and the correlation coefficients with its direct neighbors.

Refine weighted connectedness (optional): Further refine *weighted connectedness* by distinguishing between positive and negative correlations: absolute weighted connectedness (AWC) uses absolute values of correlations; positive weighted connectedness (PWC) considers only positive correlations; negative weighted connectedness (NWC) considers only negative correlations. Usually, separating positive vs. negative correlations is unnecessary for TPLoN modeling.

*Step 4*: Compute mean and variance of WC: For each species (SCN node), compute the mean (*m*) and variance (*V*) of its WC[i] vector elements (based on AWC, PWC, or NWC). This results in a (*V*, *m*) pair for each species (SCN node) in the network.

*Step 5*: Fit Taylor’s Power Law (TPL) Model (*V* = *am^b^*): Fit the TPL model (Eqn. 9) to the (*V, m*) pairs of WC [*i*], *i*=1, 2,. *N*, *N* is the number of species (nodes) in the SCN.

Finally, the interpretation of TPLoN parameters are similar with the previous TPLE ones, but the heterogeneity measured with TPLoN is in a network setting or *network heterogeneity*. Compared with TPL/TPLE, TPLoN scaling parameter (*b*) reflects heterogeneity scaling resulted from species interactions. Furthermore, randomization (permutation) tests can be applied to test the differences of TPLoN parameters between the healthy and diseased treatments.

### Species specificity and specificity diversity (SSD) framework

The SSD (species specificity and specificity diversity) framework (Ma 2024a, 2024b) is designed to achieve the following objectives:

i. *Detecting treatment-specific unique/enriched (US/ES) species* (e.g., species uniquely associated with Parkinson’s Disease (PD) patients) using the species specificity (SS) permutation (SSP) test at the species level.
ii. *Differentiating treatments holistically* (e.g., PD vs. healthy controls) using the specificity diversity (SD) and SD permutation test (SDP) at the community scale.
iii. *US/ES network (UEN) analysis*—constructing and analyzing the UEN to synthesize insights from (US or ES) species- and community-scale analyses. Optionally, the UEN can be expanded to include first-order (nearest) neighbors of US/ES species, resulting in a first-order network (FON) (Ma et al. 2024).

Compared to traditional diversity, heterogeneity, and dominance analyses, the SSD framework offers unique advantages, such as identifying treatment-specific US/ES species and integrating insights across species and community scales. While previous dominance analyses could identify dominance-enriched species (ES), they were unable to detect unique species (US). Additionally, SSD captures compositional and structural heterogeneity at regional metacommunity or global ecosystem scales through its specificity heterogeneity network (SHN), species specificity network (SSN), and the aforementioned UEN (Ma 2023, 2024a, 2024b; Ma et al. 2024).

The SSD framework comprises three core components:

i. *Species specificity (SS) and specificity permutation (SP) test*.
ii. *Specificity diversity (SD) and SD permutation (SDP) test*.
iii. *Protocols for building SHN, SSN, and UEN networks*.

Both the SP and SDP tests are based on permutation principles and rely on the metrics of SS and SD, respectively. The protocols for constructing specificity-based networks (SHN, SSN, and UEN) leverage existing complex network analysis methods, such as the previously introduced species dominance network (SDN) and SparCC networks. Below, we briefly introduce the central concepts (metrics) of SS and SD, which are essential for applying the SSD framework. For further details on the other components of SSD, refer to Ma (2024a, 2024b).

#### Species specificity (SS) and specificity permutation (SP) test

The species specificity index was first proposed by Mariadassou *et al*. (2015), who reinterpreted Dufrene & Legendre’s (1997) indicator index. Let 𝑀 = [𝑎_𝑖j_] be the OTU table representing the species abundance distributions (compositions) of microbiota, where 𝑎_𝑖j_ is the relative abundance of species *i* in sample *j*; *H* be the number of different habitats (*e.g.,* healthy and diseased treatments, here *H*=2); *S^h^* be the number of samples from habitat *h (h=1, 2, …H)*; 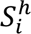 be the number of samples from habitat *h* where species *i* is present. The local species specificity (SS) index is defined as:

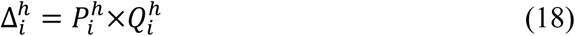

where 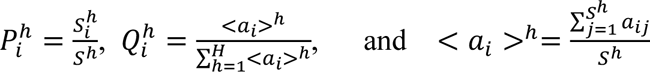

Obviously, 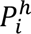 is the *prevalence* of species *i* in habitat *h*, namely, the fraction of samples from habitat *h* where species *i* was discovered. < 𝑎_𝑖_ >^ℎ^ is the average local abundances of species *i* in habitat *h*, and 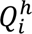 is the abundance share of habitat *h* in the total population of species *i* across all *H* habitats. Roughly speaking, the species specificity can be interpreted as the product of the species’ local (habitat or treatment specific) prevalence of distribution and its global (across all *H* habitats) abundance share.

It is worthy of noting that species specificity 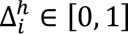, with zero indicating that the species is absent in (local) habitat *h*, with *1* indicates that the corresponding species always exists and only exists in that habitat, *i.e*., a perfect indicator species of that (local) habitat.

An extreme generalist should have 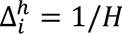, which implies that *Prevalence* 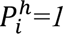, with equal abundance in all habitats. In contrast, an extreme specialist should have 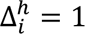, which implies that it exists in all samples of one habitat type only, but absent in all other habitats. The extreme specialist is perfect indicator species but may be rare in practice.

The specificity permutation (SP) test is designed to detect habitat (treatment) specific unique or enriched species (US or ES) based on the previous theoretical principle of SS 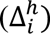 values of about specialist/generalist, guided by randomization (permutation) test principle and FDR (false discovery rate) control. 1000 times of re-sampling and *P*-value=0.05 under FDR control are adopted to implement the SP test.

#### Specificity diversity (SD) and specificity diversity permutation (SDP) test

The previous *species specificity* (SS) is defined for each species in a specific habitat type (*h*), and one can obtain a list of SS values for a group (assemblage) of species, which is specific for the metacommunity (all communities) corresponding to habitat (*h*). The list of SS in a metacommunity of habitat *h* (*h* =1, 2, …*H)* is similar to the list of species abundances in a community. However, different from species abundance, species specificity is computed for each species but across a set of communities of same habitat (metacommunity of habitat *h*), and in fact, its computation is also dependent on the alternative habitat. The similarity and difference prompted Ma (2023, 2024a, 2024b) to propose the concept of specificity diversity (SD) and defined SD as:

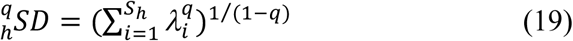

where *S_h_* is the number of species in habitat *h,* 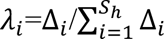 is the *relative specficity* of species *i*, *q* is the order number of specificity diversity. Obviously, the definition of SD is inspired by the definition of Hill numbers (Hill 1971) for measuring biodiversity, and both adopted the Renyi’s (1961) entropy. Obviously, the above SD metric is defined for each *metacommunity* supported by a specific habitat type *h*=1, 2, … *H*. Note that SD can also be defined for part of a metacommunity such as the assemblage of unique species in habitat *h*.

The Hill number is undefined when *q*=1, but its limit as *q* approaches to *1* exists and can be used to define SD:

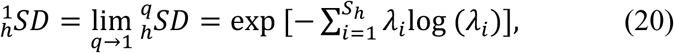

which is actually the exponential of Shannon’s entropy.

The general interpretation of specificity diversity (SD) of order *q* is that the metacommunity of habitat *h* contains 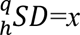 species with equivalent level of specificity. When *q*=0, SD is the number of species in the metacommunity or in the assemblage of species, such as the US specific to a treatment. SD (*q*=0) actually defaults to species richness in the metacommunity or assemblage. When *q*=1, SD (*q*=1) represents the number of species with common level of specificity. When *q*=2, SD (*q*=2) represents the number of species with dominant (higher) level of specificity. Although the relationship between generalist-specialist spectrum with species specificity is complex as discussed in Ma (2024a), larger specificity suggests that the species is more likely being a specialist and less likely being a generalist. Therefore, SD at higher diversity order (*q*) captures the diversity (number) of more specialist species, and SD at lower diversity order (*q*) captures the diversity of more generalist species. When *q*=0, specificity is not weighted in the computation of SD, and the SD simply defaults to the species richness.

Given that specificity diversity (SD) measures the diversity (in equivalent numbers) of species across different positions on the generalist-specialist spectrum, the SD permutation (SDP) test can be employed to assess shifts in this spectrum caused by treatment effects, such as disease. The SDP test is designed to rigorously compare the SD of two species assemblages (at the community or metacommunity level) under different treatments, such as healthy and diseased cohorts, using a statistically robust framework (Ma 2024a).

### Neural Networks and Machine Learning for Diagnosing NNP Disorders

To explore the automatic classification of gut microbiome samples into their respective treatment groups—healthy controls or those diagnosed with NNP disorders, including Alzheimer’s disease (AD), autism spectrum disorder (ASD), mood disorders (MD), and Parkinson’s disease (PD)—we employed six machine learning models. These models, trained with the OTU abundance tables derived from gut microbiomes across 23 case studies, included: (i) Deep Neural, (ii) Random Forest, (iii) Support Vector Machine, (iv) Logistic Regression, (v) Gradient Boosting, and (vi) K-Nearest Neighbor.

Each model was trained on 80% of the datasets, with the remaining 20% reserved for testing. Furthermore, to address sample heterogeneity, the process was repeated 100 times, generating 100 precision values for each model. To explore optimal strategy for differentiate (diagnose) NNP disorders, we designed the following five modeling/testing schemes:

i. Scheme I: Comparisons between the healthy cohort and each of the four disease cohorts individually: Healthy *vs*. AD; Healthy *vs*. ASD; Healthy *vs.* MD; Healthy *vs*. PD, a total of 4 sets (or 4*6=24) of AI/machine learning models.
ii. Scheme II: Pairwise comparisons among the four disease cohorts: AD *vs*. ASD; AD *vs*. MD; AD *vs*. PD; ASD *vs*. MD; ASD *vs*. PD; MD *vs.* PD, a total of 6 sets (or 6*6=36) models.
iii. Scheme III: (A) Simultaneous classification of the four NNP disorders AD vs. ASD vs. MD vs. PD; (B) Simultaneous classification of the four NNP disorders and healthy control AD vs. ASD vs. MD vs. PD vs. Healthy. The distinction between Scheme III-A and III-B lies in whether the healthy control is excluded or included.
iv. Scheme IV: Select species with significant differences in species specificity (SS), specifically those identified as unique species (US) and enriched species (ES), excluding species without significant SS differences. Apply Scheme I and Scheme II using the six selected AI machine learning algorithms.
v. Scheme V: (A) Rank all species by species specificity (SS) in descending order and select the top 50 species with the highest SS. Apply Schemes I, II, and III using the six selected AI/machine learning algorithms. (B) Additionally, this scheme includes training and testing with the top 25 species (those with the highest SS) to apply Schemes I-III.

This structured approach ensures a comprehensive evaluation of the models’ performance across various diagnostic scenarios, offering robust insights into their ability to distinguish between healthy and diseased states, as well as among specific NND disorders. In particular, the last two schemes are designed to test the hypothesis that “less can be more”—using SSD (species specificity and specificity diversity) to select the most informative (“specific”) species for diagnosis.

## Results

### Diversity Analysis

Four types of diversity analyses—alpha-diversity, beta-diversity (Anna Karenina Principle), gamma-diversity (diversity scaling and potential diversity), and network diversity (ND)—were applied to analyze the five treatment groups: Alzheimer’s disease (AD), autism spectrum disorder (ASD), mood disorders (MD, including depression and bipolar disorder), Parkinson’s disease (PD), and the healthy control.

#### Alpha-Diversity in Hill Numbers

Table S2 presents the mean and standard error of microbiome diversity in Hill numbers for four NNP (Neurodegenerative, Neurodevelopmental, and Psychiatric) disorders—ASD, MD, AD, and PD—derived from 23 published case studies (see Table S1 for brief descriptions). Table S3 displays the results of nonparametric two-way ANOVA and post-hoc multiple comparisons using the Scheirer-Ray-Hare test from the R-companion package in R. These results highlight the effects of Disease Type (DT), Health Status (HS), and their interaction on diversity. All tests were conducted at a significance level of *P-*value=0.05.

Both Disease Type (DT) and Health Status (HS) significantly influenced diversity across all diversity orders (*q*=0-2) (*P*-value < 0.001 for both DT and HS). The interaction effects between DT and HS were not significant, with *P*-values ranging from 0.187 to 0.250 (Fig 2A).

**Fig 2A.**
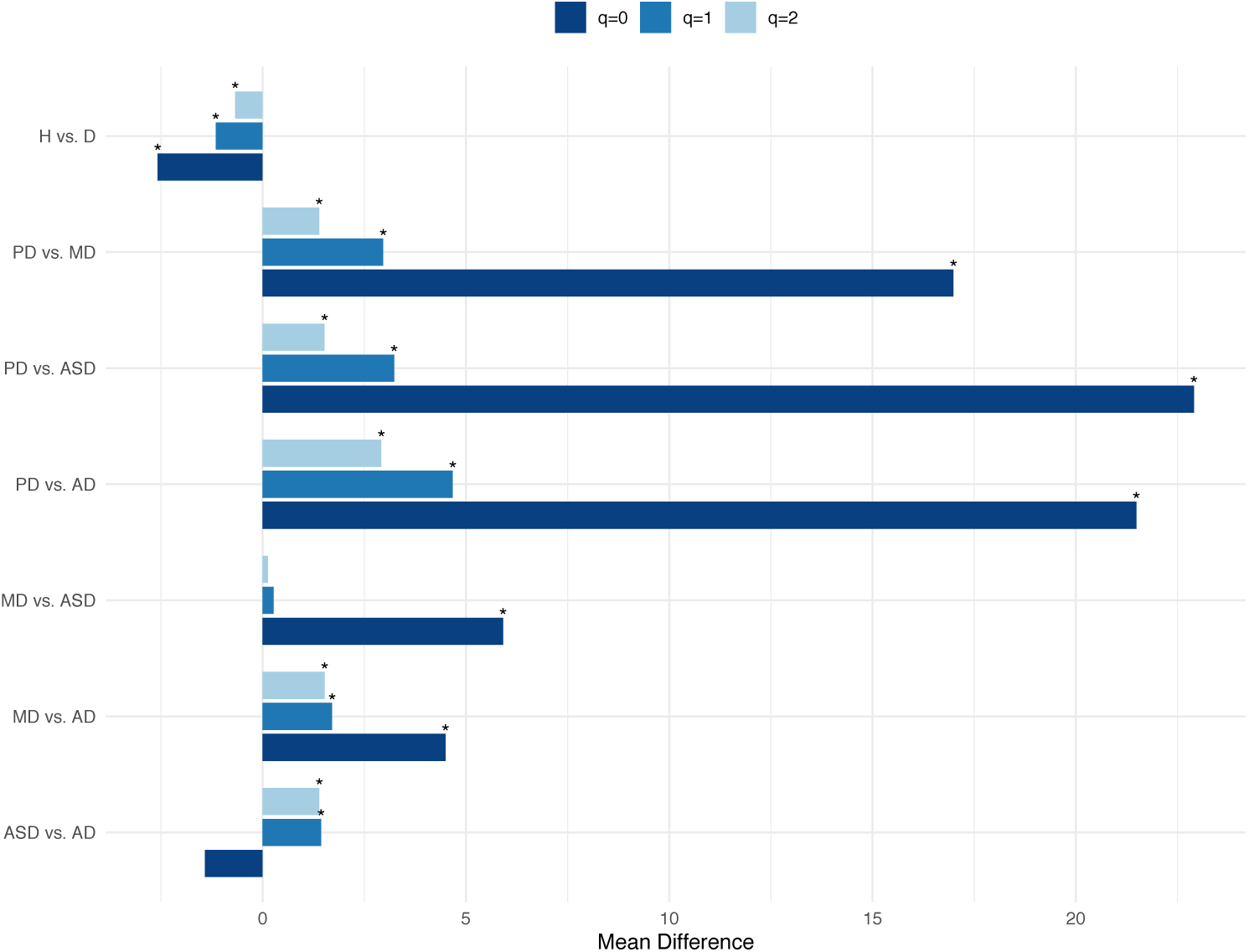
Significant difference tests for comparing the alpha-diversity (at different diversity orders *q*=0, 1, 2): Asterisks (*) indicate significant differences with *p* < 0.05 for ANOVA. (Abbreviations: H= Healthy; D=Diseased).

Post-hoc multiple comparisons (Table S4) revealed significant differences in 83% (5 out of 6) of pairwise comparisons between the four DTs. The only non-significant pair was MD *vs*. ASD at *q* =1 and *q*=2 (*P*-value = 0.800-0.913). However, this pair of MD *vs*. ASD showed significant differences at *q*=0 (species richness) with P-value < 0.001 (Fig 2A).

Regarding Health Status (HS), diseased treatments consistently exhibited higher alpha-diversity than the healthy controls across all diversity orders (*q*=0-2) (Fig 2A).

Overall, PD patients displayed the highest alpha-diversity, while ASD patients showed the lowest. Specifically,

i. At q=0: PD > MD > AD; PD > MD > ASD.
ii. At q=1: PD > ASD > AD; PD > MD > ASD.
iii. At q=2: PD > MD > AD; ASD > AD; PD > ASD.

#### Anna Karenina Principle (AKP) Tests Based on Beta-Diversity

Tables S5 and S6 present the mean beta-diversity and AKP test results based on beta-diversity. Diseased treatments exhibited significantly higher (*P*-value<0.001) beta-diversity across all four DTs, consistent with the AKP. Among the four DTs, the order of beta-diversity was: MD > PD > AD > ASD (Fig 2B).

**Fig 2B.**
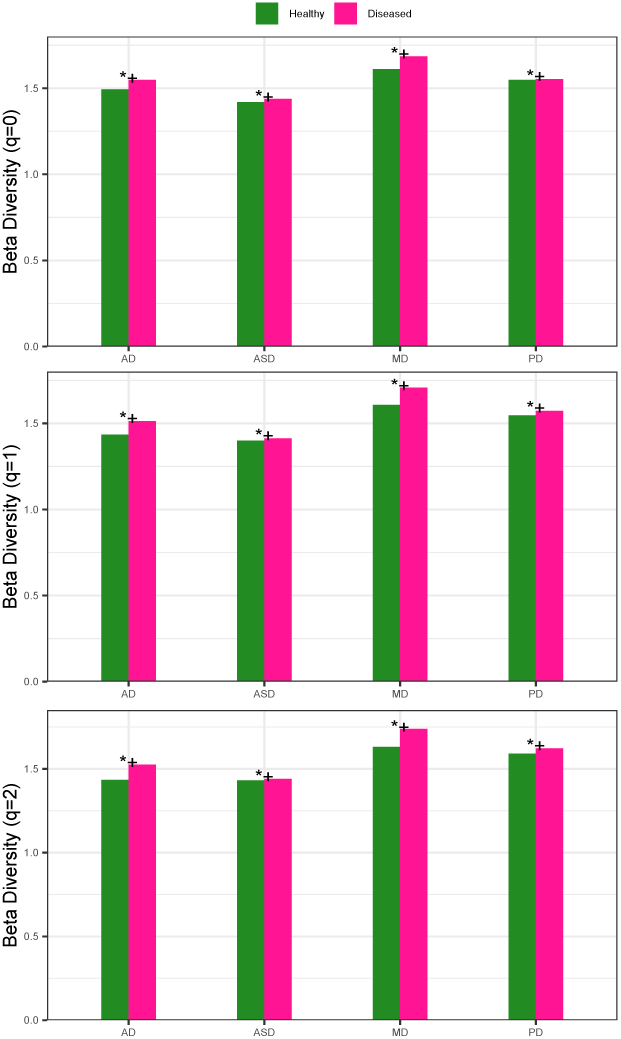
The AKP (Anna Karenina Principle) test at various diversity order *q*=0, 1, & 2. Legends: ‘*+’ for AKP; ‘*-’ for Anti-AKP; empty (no marking) for non-AKP. All tested comparisons of the four NNP disorders indicate AKP effects.

#### Diversity Scaling and Potential Diversity Analyses

Table S7 shows the fitting of DAR (diversity-area relationship) models (with 1,000 random permutations of microbiome samples for robustness) for four NNP disorders—AD, ASD, MD and PD—from 23 published case studies, along with the healthy control. Table S8 presents the results (*P*-values and percentages of significant differences) from permutation tests for DAR parameters. The permutation tests revealed that the DAR scaling parameter (*z*) remained invariant across disease types at all diversity orders (*q*=0-2). However, potential diversity (*D*_max_) varied across DTs and diversity orders (Fig 2C):

i. At *q*=0: *D*_max_ differed in 10% (1 out of 10) of comparisons.
ii. At *q*=1: D_max_ differed in 90% (9 out of 10) of comparisons.
iii. At *q*=2: D_max_ differed in 60% (6 out of 10) of comparisons.

**Fig 2C.**
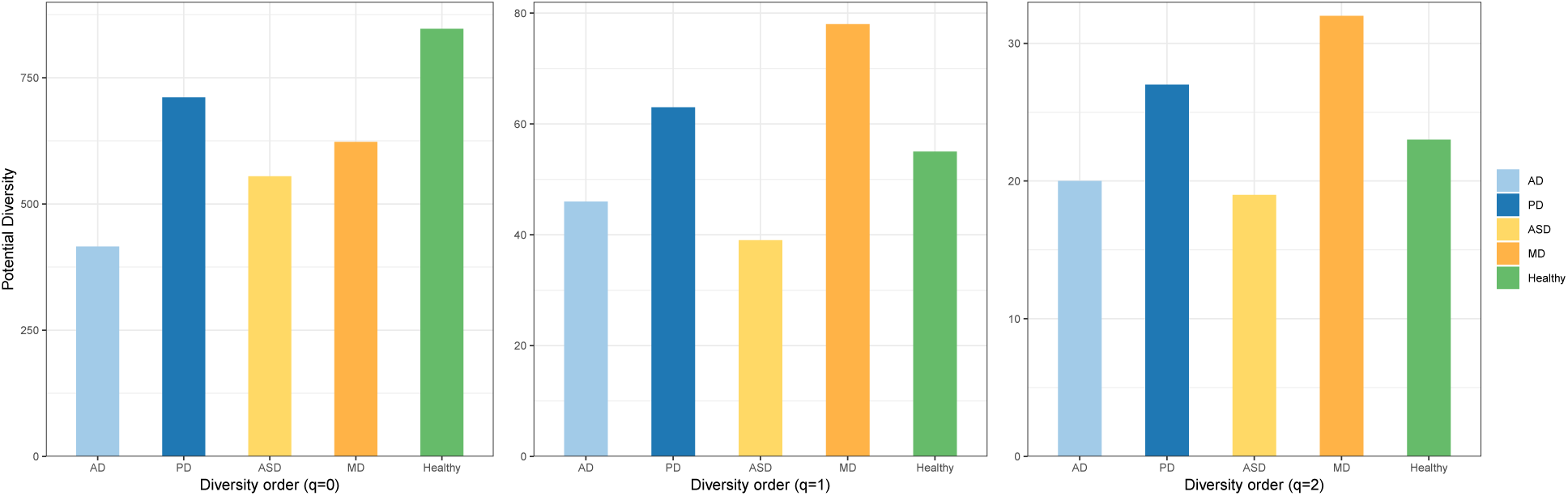
The potential microbial diversity (*D_max_*) at different diversity order (*q*=0, 1, 2) for the four NNP disorders and healthy control, respectively

These results suggest that potential diversity measured as species richness (*q*=0) is generally consistent across disease types and health status. However, the potential diversity of common species (*q*=1) and dominant species (*q*=2) show significant variability, particularly at *q*=1.

#### Network Diversity (ND) Analysis

We first build species co-occurrence network (SCN) by using the computing protocols for species dominance networks (SDNs) (Ma & Ellison 2019). Spearman’s correlation coefficients were applied to species dominances for building the SCNs (*i.e.,* SDNs) of the four NNP disorders and the healthy control (FDR control with *P*-value<0.05 was applied).

Table S9 lists the three types of network diversity (ND) for each treatment (AD, ASD, MD, PD, and healthy controls): (i) ND of species relative abundances (NDSA), (ii) ND of link probability (NDLP), (iii) ND of link abundances (NDLA). Table S10 provides the *P*-values from permutation tests for NDs, and Fig 2D illustrates the NDs of the five treatments in bar charts.

**Fig 2D.**
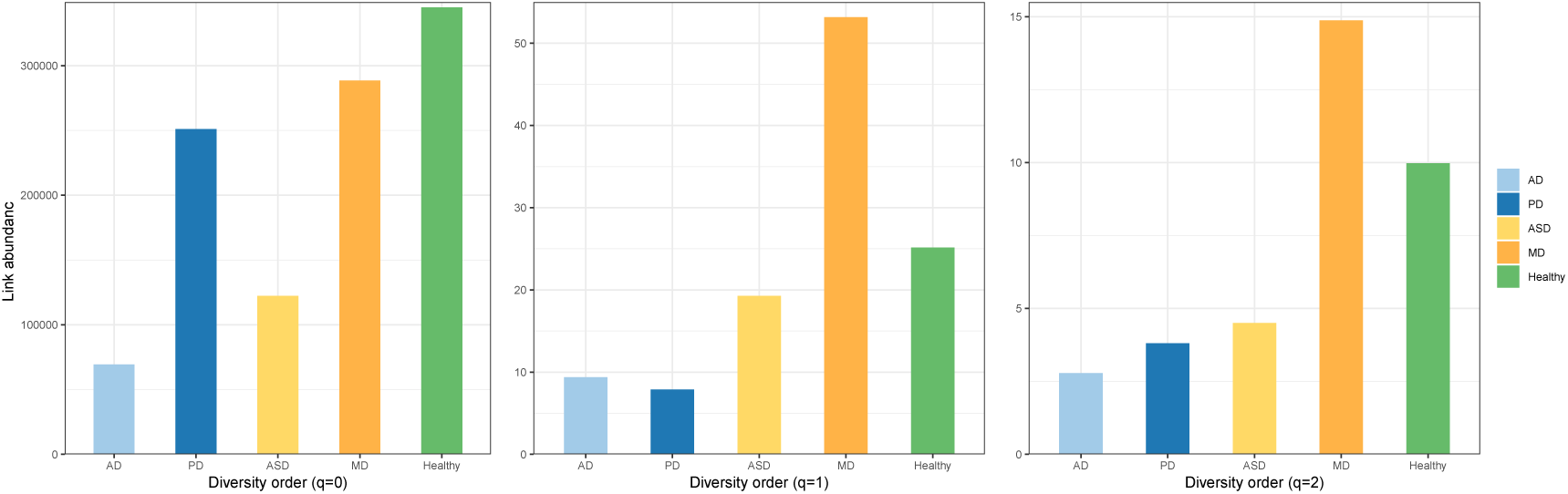
The NDLA (network diversity of link abundance) of the gut microbiomes associated with the NNP disorders

From a network diversity type perspective:

i. NDSA: 70-90% of comparisons showed significant differences (70% at q=0; 90% at q=1-2).
ii. NDLP: 0-70% of comparisons showed significant differences (70% at q=0; 0% at q=1-2).
iii. NDLA: 20-70% of comparisons showed significant differences (70% at q=0; 20-30% at q=1-2).

From a diversity order perspective:

i. At q=0: all three NDs showed significant differences in 70% (7 out of 10) of comparisons.
ii. At q=1: NDSA showed 90% differences, NDLP showed 0%, and NDLA showed 30%.
iii. At q=2: NDSA showed 90% differences, NDLP showed 0%, and NDLA showed 20%.

These findings highlight the variability in network diversity across treatments and diversity orders, with NDSA demonstrating the highest consistency in detecting differences across all diversity orders. Among the three types of NDs, NDSA primarily captures the diversity of network nodes, while NDLP and NDLA capture the diversity of network links. At higher diversity orders (q > 1), network diversity, particularly NDLP and NDLA, approaches network heterogeneity due to their focus on species links (interactions).

With this context, it is evident that the patterns revealed by NDSA align largely with previous findings from classic diversity metrics. The 70%-90% differences observed here are consistent with the 83% differences (Table S4) found in traditional alpha-diversity analysis. In contrast, what NDLP and NDLA reveal—especially at higher diversity orders (*q*=1-2)—is more about heterogeneity than diversity. The 0%-30% differences observed align with the findings in the heterogeneity section below. Notably, NDLA measures a combination of species abundances and species interactions, while NDLP largely serves as a pure proxy for species interactions. In other words, NDLA can be considered a hybrid proxy of both diversity and heterogeneity, while NDLP is a pure proxy of heterogeneity. This distinction explains why NDLP shows 0% differences among disease types for *q*>0. We hypothesize that heterogeneity primarily operates on an evolutionary time scale, while diversity reflects ecological time scales. Therefore, heterogeneity scaling (or alterations) should be less pronounced than diversity over the time scale in which these observations were made, including during the course of this research. Further discussion on this topic is deferred to later sections.

##### Heterogeneity Analysis

Three types of heterogeneity analyses—dominance analysis, Taylor’s Power Law Extensions (TPLE), and Taylor’s Power Law of Networks (TPLoN)—were applied to analyze the five treatment groups: Alzheimer’s disease (AD), autism spectrum disorder (ASD), mood disorders (MD), Parkinson’s disease (PD), and healthy controls.

#### Dominance Analysis

Tables S11 and S12, along with Fig 3, present the results of community dominance analysis supported by two-way ANOVA and post-hoc multiple comparisons. Both Disease Type (DT) and Health Status (HS), as well as their interactions, were found to be statistically significant (*P*-value < 0.001) across all three aspects. Specifically,

i. Disease Types (DTs): Post-hoc pairwise comparisons revealed significant differences in 66.7% of cases. The remaining 33.3% of non-significant differences were attributed to ASD vs. AD and PD vs. MD.
ii. Health Status (HS): Diseased treatments exhibited significantly higher community dominance compared to healthy controls.
iii. Interaction Effects (DT x HS): 78.6% of multiple comparisons showed significant differences in community dominance.

**Fig 3.**
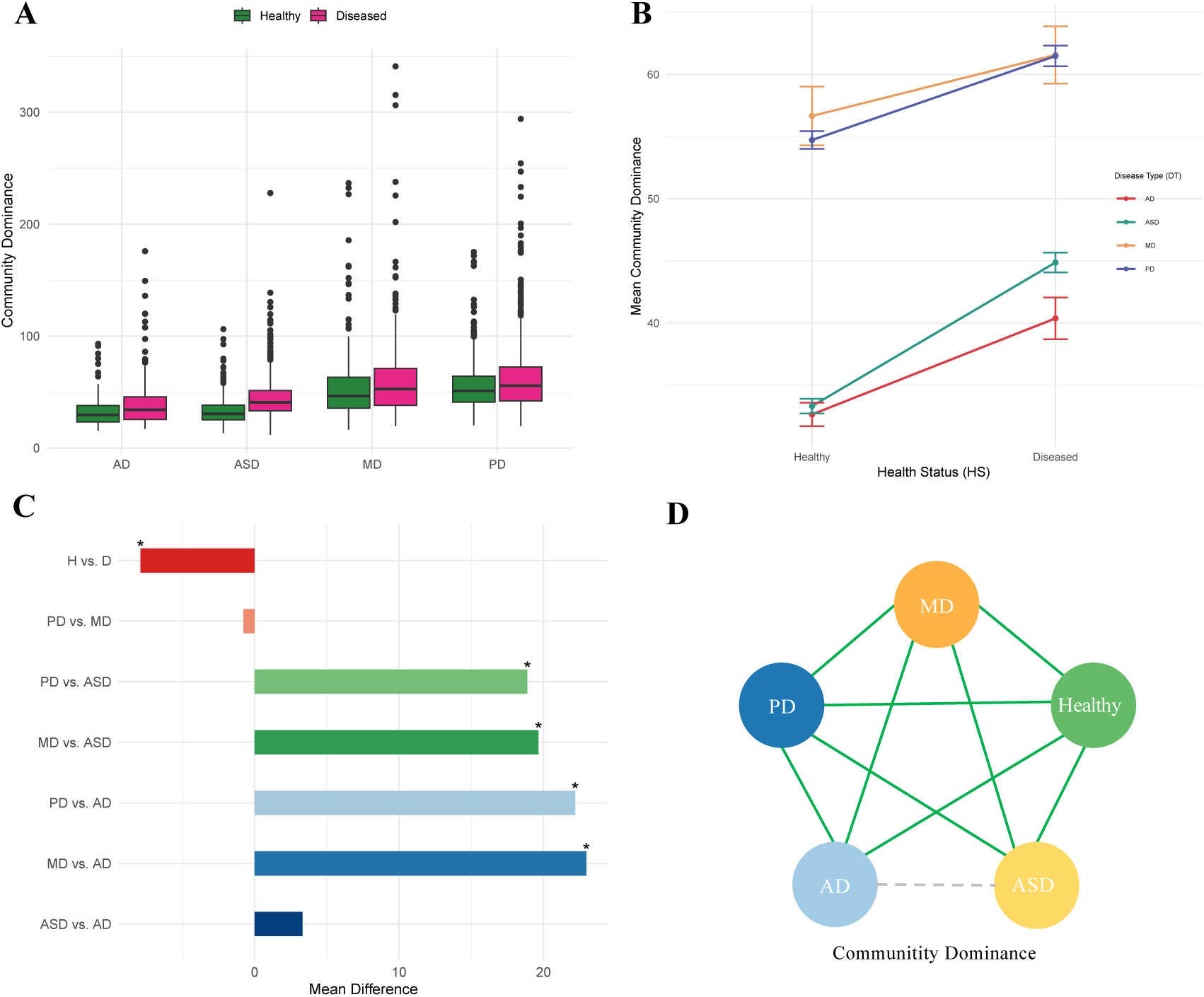
Dominance analysis via two-way ANOVA, the effects of both disease type (DT) and healthy status (HS) as well as their interactions are statistically significant; (**A)** Standard bar charts showing the community dominance: the dominance order is MD>PD>ASD>AD and D>H; **(B**) Interaction between NNP disease type (DT) and health status (HS) on Community Dominance. **(C)** Mean difference for the comparison of community dominance (Asterisks indicate *p* < 0.05 for ANOVA) Abbreviations: H= Healthy; D=Diseased. **(D)** Pairwise comparisons between health and 4 NNP disorder types. The green line indicates a significant difference between the two groups and the grey dotted line indicates no significant difference between the two groups.

#### Taylor’s Power Law Extensions (TPLE) Analysis

Table S13 provides the TPLE parameters for each of the five treatments (AD, ASD, MD, PD, and healthy controls, while Table S14 presents the PP-values from permutation tests comparing these parameters. Fig 4A illustrates the fitting of Type-I TPLE for measuring community spatial heterogeneity (CSH), and Fig 4B displays the fitting of Type-III TPLE for assessing mixed-species population aggregation (MSA). Specifically,

i. *Type-I TPLE (Community Spatial Heterogeneity)*: The order of the scaling parameter *b* was: MD (*b*=2.242) > ASD (1.956) > *PD (1.884) > Healthy (1.878)* > AD (1.566). No significant differences in *b* were found between healthy controls and PD, but significant differences were detected in all other pairwise comparisons (9 out of 10). The ln(*a*) parameter showed differences in 100% of comparisons, while the HCT (heterogeneity critical threshold) parameter exhibited differences in 70% of cases.
ii. *Type-III TPLE (Mixed-Species Population Aggregation):* Type-III TPLE modeling revealed much smaller differences between treatments, as evidenced by the narrow range of parameters. The scaling parameter *b* ranged from 1.599 (healthy) to 1.671 (ASD), with significant differences only detected between ASD and PD. The ln(*a*) parameter showed significant differences in 40% of comparisons, and the HCT parameter in 10% of cases.

**Fig 4A.**
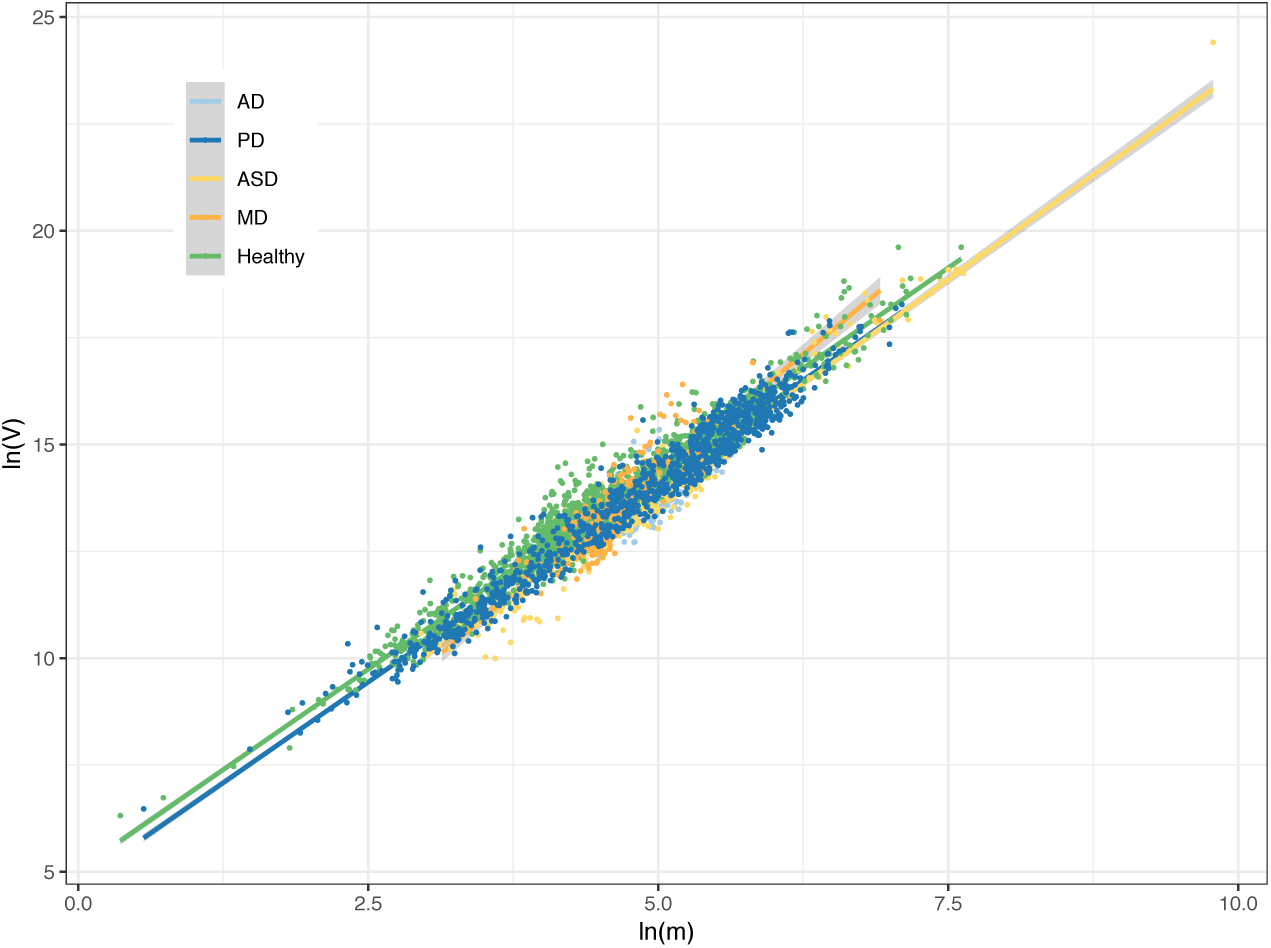
Fitting Type-I TPLE (Taylor’s power law extension) for measuring community spatial heterogeneity: MD (major depression) showed the highest TPLE scaling parameter b=2.242, while AD (Alzheimer disease) showed lowest (*b*=1.566).

**Fig 4B.**
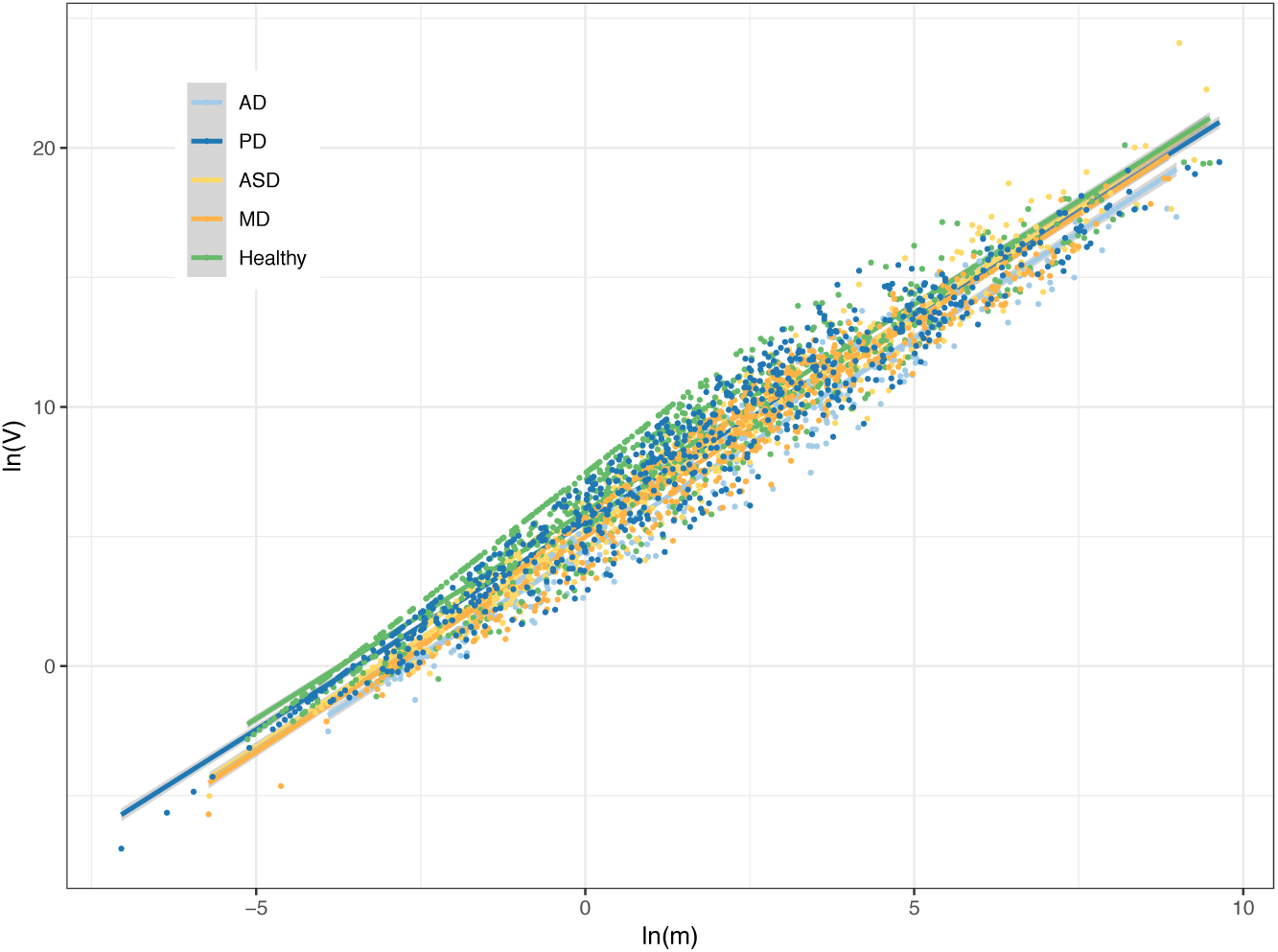
Fitting Type-III TPLE (Taylor’s power law extension) for measuring community spatial heterogeneity: ASD (autism disorder) showed the highest TPLE scaling parameter *b*=1.671, while the healthy showed lowest (*b*=1.599). TPLE-b showed significant differences in only 10% (1 out of 10) comparisons (ASD *vs*. PD). ln(a) in 40% and HCT in 10% of the comparisons.

The contrasting patterns between Type-I TPLE (community spatial heterogeneity) and Type-III TPLE (mixed-species population aggregation) suggest that heterogeneity at the community (or meta-community) scale is less stable than at the population scale. This may reflect the simple additive effects of species-level heterogeneity that emerge at the community or meta-community level.

#### Taylor’s Power Law of Networks (TPLoN)

The species dominance networks (SDNs), based on species dominance and Spearman’s correlation coefficients, were used for TPLoN modeling. These are the same SDNs previously built for computing the network diversity (ND) previously. Table S15 lists the TPLoN parameters for the five treatments, and Table S16 provides the *P*-values from permutation tests comparing these treatments.

i. The order of the scaling TPLoN parameter *b* was: *Healthy (1.954) > PD (1.919)* > ASD (1.909) > MD (1.838) > AD (1.801).
ii. Significant differences in TPLoN-*b* were detected in 90% of pairwise comparisons, with the only non-significant difference observed between healthy controls and PD. This finding is consistent with previous results from permutation tests of the TPLE scaling parameter (*b*), where differences were also observed in 90% of cases, or 9 out of 10 instances.
iii. The ln(*a*) parameter exhibited differences in 100% of comparisons, while the HCT parameter (*m*_0_) showed significant differences in 70% of cases.

Fig 4C illustrates the fitting of TPLoN models, and Fig 4D displays the model parameters for the various treatments.

**Fig 4C.**
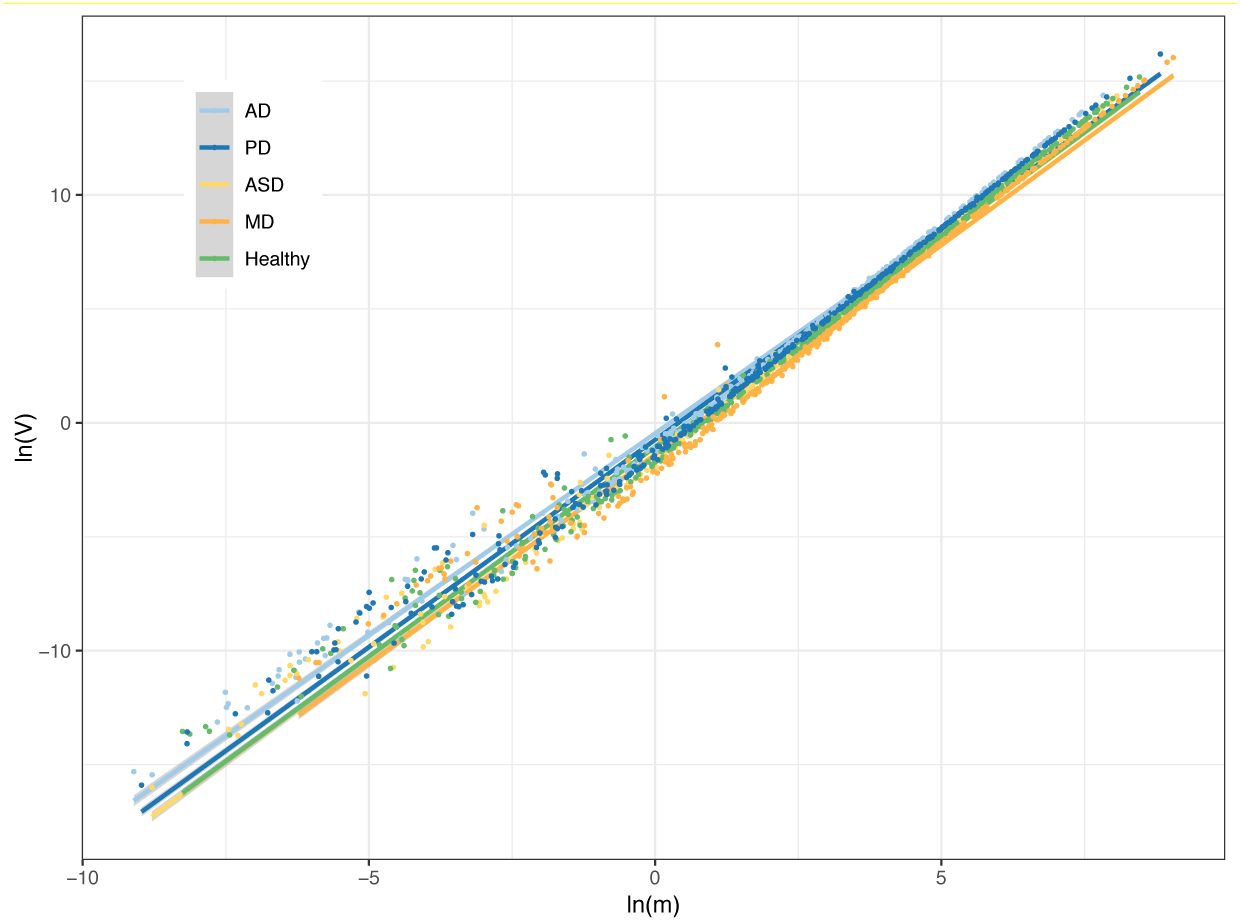
Fitting the TPLoN (Taylor’s power law of network) model for measuring network heterogeneity of the gut microbiomes associated with NNP disorders

**Fig 4D.**
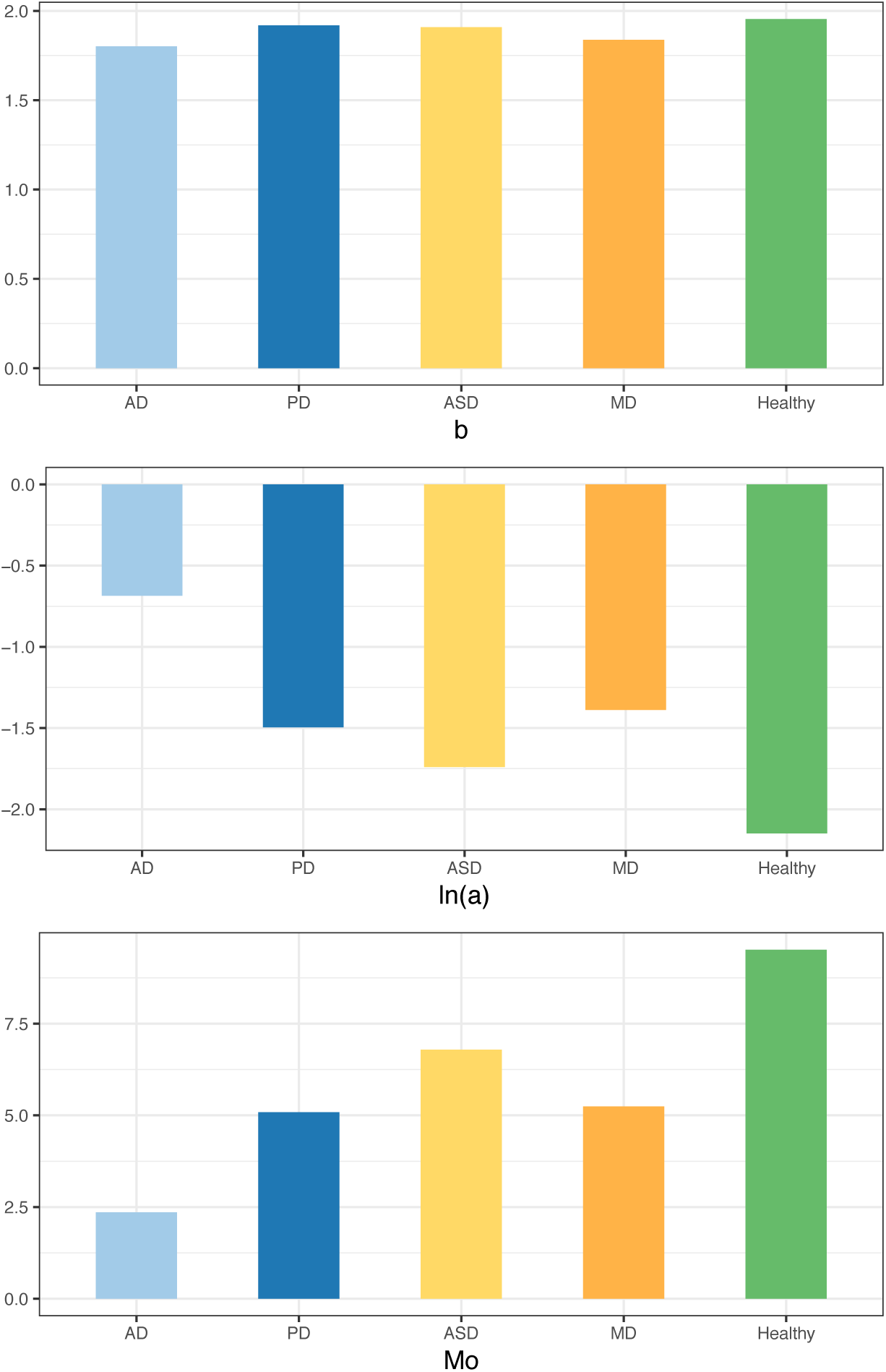
The network heterogeneity parameters of TPLoN (Taylor’s power law of network) From top to bottom, *b*, ln(*a*), and *M0* (Heterogeneity Critical Threshold). Healthy group has the highest scaling parameter (b=1.954), and AD has the lowest (b=1.801); significant differences were found in 90% comparisons. ln(a) exhibited significant differences in all 10 comparisons. HCT exhibited significant differences in 70% (7 out of 10) comparisons.

### SSD (species specificity and specificity diversity) Analysis

#### Species Specificity (SS) and Specificity Permutation (SP) Tests

Tables S17A-S17J (in MS-Excel sheets) tabulate the unique species (US) and enriched species (ES) identified for each treatment based on specificity permutation (SP) tests with FDR control of *P-*value<0.05. Fig 5A illustrates the distribution of US/ES species in AD-associated microbiomes using volcano plots. Unlike the community- or meta-community-level analyses presented earlier, the specificity permutation (SP) tests within the SSD framework provide species-level insights. Specifically, the US/ES lists offer potential candidates for diagnostic biomarkers and/or therapeutic probiotics for NNP disorders.

**Fig 5A.**
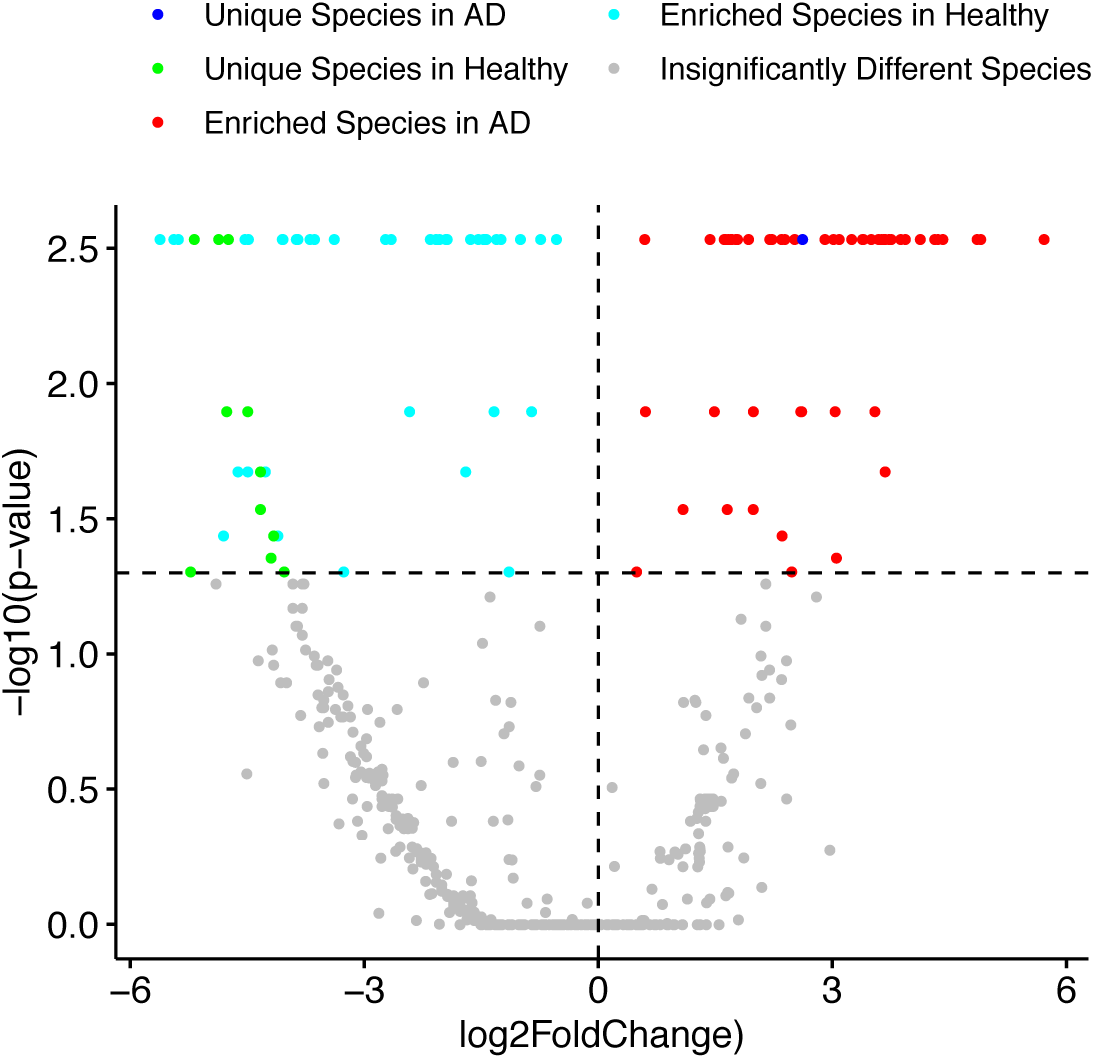
The volcano plot depicts the distribution of US (unique species) and ES (enriched species) in AD patients and healthy controls across five regions (With FDR and *P*<0.05). The lower section illustrates species with no significant difference in specificity between the two groups. The upper left region displays the US/ES species in healthy controls, while the upper right region represents those in AD patients.

Fig 5B presents the network graph of uniquely present/enriched or depleted (US/ES) microbial species in Alzheimer’s disease (AD) patients. The left block of the graph represents US/ES species in the healthy control group (equivalently, the species depleted in AD patients), while the right block depicts US/ES species in AD patients (which are depleted in the healthy control). Fig 5C displays the corresponding network graph for US/ES microbial species in mood disorders (MD) patients, with the left and right blocks interpreted similarly to those in Fig 5B for AD. Notably, the network graphs for AD (Fig. 5B) and MD (Fig. 5C) exhibit distinct structural denseness/sparsity, even upon visual inspection.

**Fig 5B.**
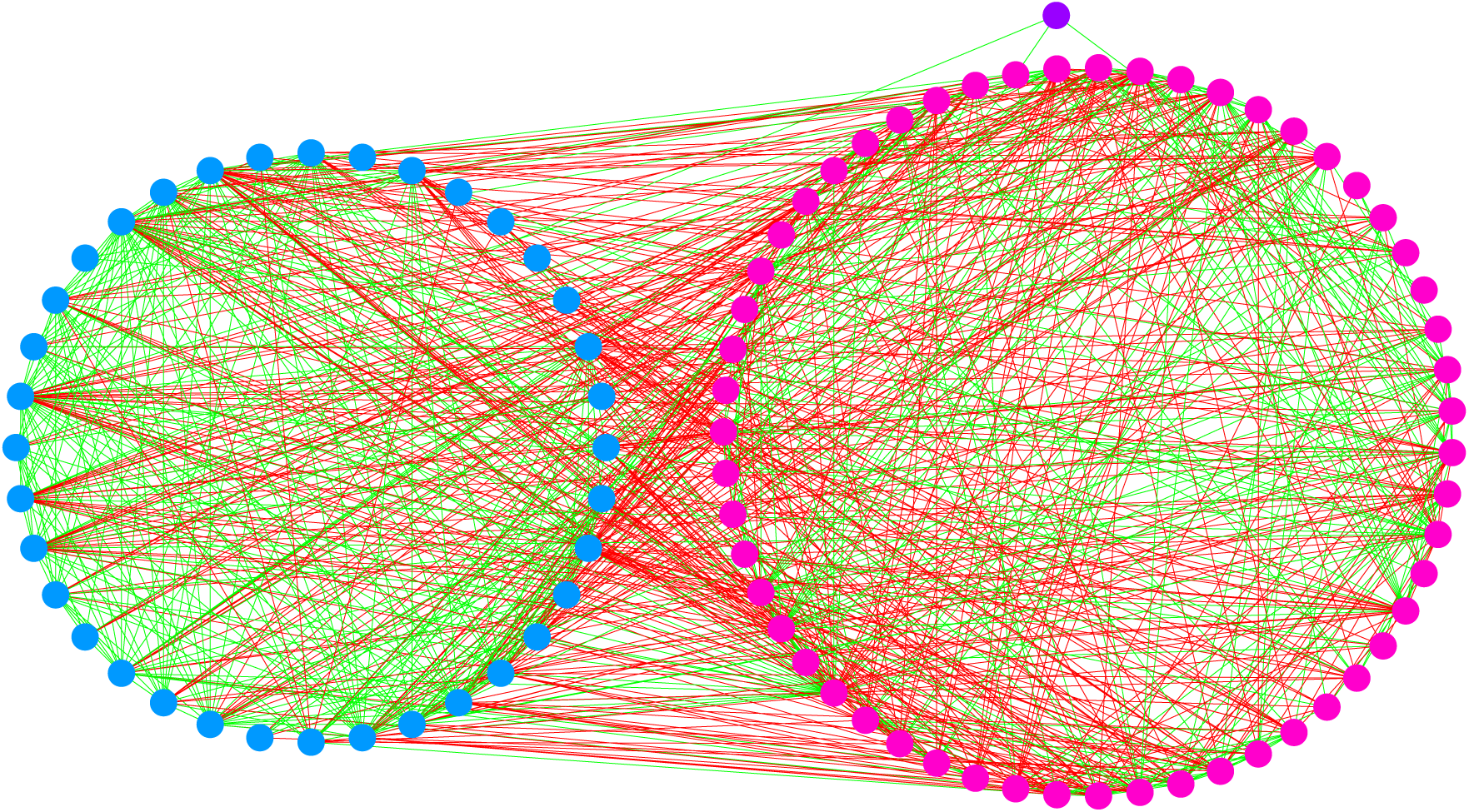
Network graph of the US (unique species) / ES (enriched species) in the gut microbiome of Alzheimer’s disease (AD), illustrating interactions (strictly co-occurrences) among US/ES groups. Correlations were computed using the SparCC algorithm with a false discovery rate (FDR) of *P* < 0.05. Nodes are color-coded as follows: pink for species enriched in AD (ESD), blue for species enriched in healthy controls (ESH), and purple for species unique to AD (USD). Green lines represent positive correlations, while red lines indicate negative correlations. Network density is defined as the number of edges divided by the number of nodes. The P/N ratio is calculated as the number of positive edges divided by the number of negative edges. A higher P/N ratio indicates a lower proportion of negative edges in the network and implies a stronger cooperative network. P/N Ratio: ESH (29.571) > ESD (1.656) > Inter-Group (0.244) (Inter-Group of ESH and ESD) Network: Density: ESH (6.294) > ESD (6.182) > Inter-group (4.284)

**Fig 5C.**
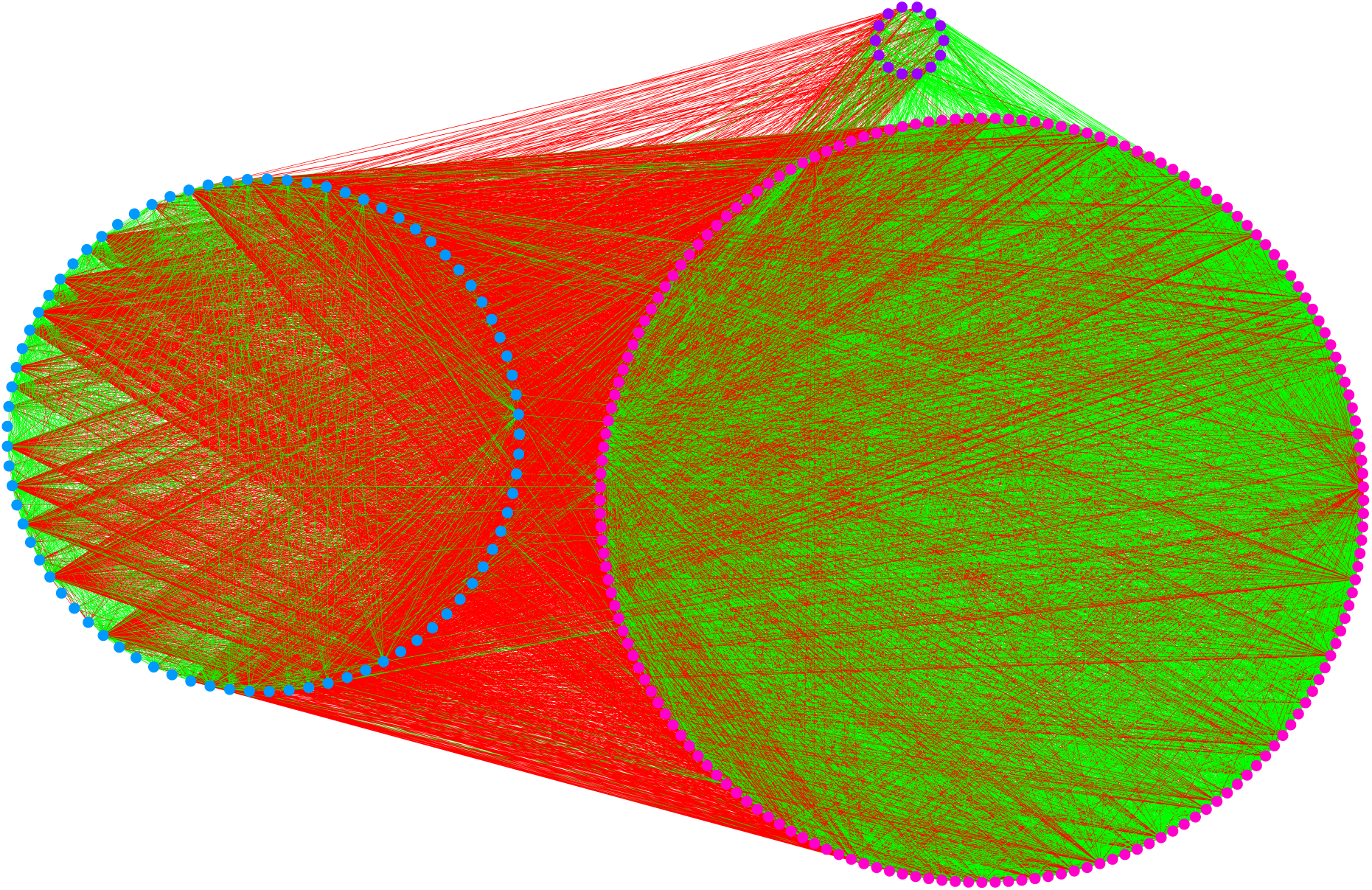
Network graph of the US (unique species) / ES (enriched species) in the gut microbiome of MD, illustrating interactions (strictly co-occurrences) among US/ES groups. Correlations were computed using the SparCC algorithm with a false discovery rate (FDR) of *P* < 0.05. Nodes are color-coded as follows: pink for species enriched in MD (ESD), blue for species enriched in healthy controls (ESH), and purple for species unique to MD (USD). Green lines represent positive correlations, while red lines indicate negative correlations. Network density is defined as the number of edges divided by the number of nodes. The P/N ratio is calculated as the number of positive edges divided by the number of negative edges. A higher P/N ratio indicates a lower proportion of negative edges in the network and implies a stronger cooperative network. P/N Ratio: ESH (8.429) > ESD (4.568) > Inter-group (0.144) Density: ESD (36.022) > Inter-group (20.441) > ESH (13.7141) The correlations within the USD group are all positive. P/N Ratio: Inter-group (USD *us*. ESD) = 8.059 (Density=2.943) P/N Ratio: Inter-group (USD *us*. ESH) = 0.030 (Density=4.556) More negative correlations between USD and ESH (lower P/N ratio) than between USD & ESD

Fig 5B and 5C also display species interactions within and between US/ES species. We use two network metrics, network density (number of links divided by node) and P/N ratio (the ratio of positive to negative correlations) to reveal their interaction patterns. In Fig. 5B for AD, the P/N ratio for enriched species (ES) in the healthy control group (ESH) is 29.571, significantly higher than that of the diseased treatment group (AD) or ESD (1.656), with both values substantially exceeding the inter-group P/N ratio (0.244) between ESH and ESD. Since higher P/N ratios indicate fewer negative correlations (a proxy for interactions), these ratios demonstrate that the enriched species (ESs) in the healthy network are significantly more harmonious (cooperative) than those in the AD network, and both groups exhibit greater cooperation compared to the interactions between the two groups. In other words, within group interactions (either within ESH or ESD) are more cooperative than between group interactions (between ESH and ESD). This pattern arguably highlights the most conspicuous impacts of AD disorder on the gut microbiomes.

Similarly, in Fig 5C for MD, The P/N ratios reveal that the enriched species (ESs) in the healthy network (ESH = 8.429) are significantly more harmonious (cooperative) than those in the MD network (ESD = 4.568), and both groups exhibit greater cooperation compared to the interactions between the two groups (Inter-group = 0.144). This suggests a more balanced and cooperative microbial community in the healthy group, while the MD group shows reduced harmony, and the interactions between healthy and MD groups are the least cooperative. These results demonstrated by Fig 5B and 5C are also supported by previous AKP tests.

From a network density perspective, the network densities for the three blocks (groups) in AD are ESH (6.294) > ESD (6.182) > Inter-group (4.284). In contrast, the corresponding values for MD are ESD (36.022) > Inter-group (20.441) > ESH (13.714). The 3- to 6-fold differences in network density between the two diseases highlight another significant distinction between AD and MD, which is further elaborated on below. MD exhibits significantly denser species interactions compared to AD.

Fig 5D (see also Table S18) illustrates the specificity correlation relationships among the five treatments (4 NNP disorders and healthy control) in a pentagram format. In Table S17K (in MS-Excel sheet), we listed the species specificity of each species in each NNP disorder.

**Fig 5D.**
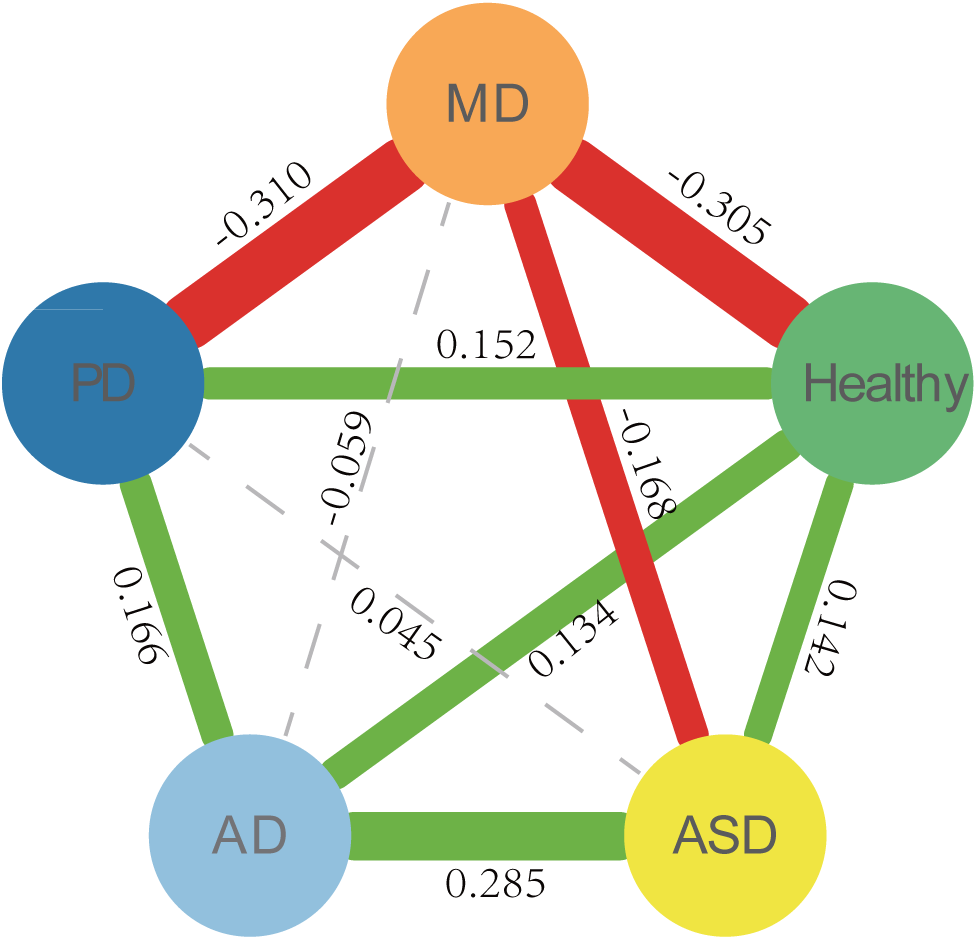
The specificity correlations between five groups (four NNP disorders and healthy control). Correlation coefficients were calculated using Spearman with FDR control (*P*-value=0.05). The green line indicates a positive correlation, the red line indicates a negative correlation, and the thickness of the line indicates the strength of the correlation. The grey, dashed lines indicate no significant correlations.

Key observations include:

i. MD (Mood Disorders): Exhibited negative correlations with ASD, PD, and healthy controls, and no significant correlation with AD. This highlights the unique specificity of MD, suggesting that its microbiome profile is distinct from both other neurological disorders (neurodegenerative and neurodevelopmental disorders) and healthy controls.
ii. Healthy Controls: Showed negative correlations with MD but positive correlations with AD, ASD, and PD. This reinforces the distinctiveness of MD compared to other disorders and healthy controls.
iii. PD (Parkinson’s Disease) and AD (Alzheimer’s Disease): Positively correlated in their specificity. Both PD and AD showed positive correlations with healthy controls, indicating that the dysbiosis associated with these diseases is not so severe as to completely differentiate them from healthy states.
iv. PD and ASD (Autism Spectrum Disorder): No significant correlation in specificity, possibly due to age differences between ASD and PD patients.
v. MD and AD: No significant correlation, but MD showed a negative correlation with PD. This suggests that the dysbiosis associated with AD is less pronounced than that associated with PD. This observation is supported by previous heterogeneity analyses: In Type-I TPLE modeling, the scaling parameter *b*-values were 1.884 (PD) *vs.* 1.566 (AD). In TPLoN modeling, the *b*-values were 1.919 (PD) vs. 1.801 (AD). These differences were statistically significant (*P*-value < 0.001). That is, although both PD and AD are neurodegenerative disorders, their heterogeneity scaling degrees (*b*) differ significantly.

#### Specificity Diversity (SD) and SD Permutation Tests

Tables S19-S22 present the specificity diversity (SD) and SD permutation (SDP) test results. Specificity diversity (SD), as defined in Eq. (19), summarizes the species specificity (SS) values of species within a given species assemblage, such as the unique species (US) of PD patients. While SD uses the same Rényi entropy formula as Hill numbers for defining biodiversity metrics, its interpretation differs. Traditional species diversity in Hill numbers summarize species abundance information within a community, whereas SD of SSD framework summarizes species specificity (Ma 2024a, 2024b).

Before summarizing the SDP test results, it is important to clarify two subtle technical points regarding the designation of species assemblages (groups) for computing and testing SD and regarding the diversity order (*q*=0). The SDP test compares the SD of two species assemblages contained in two separate treatments. For example: Table S19A displays the SDP test results for the SD of USs in the “former” treatment (*e.g*., healthy controls). Table S19B displays the SDP test results for the SD of USs in the “latter” treatment (*e.g.,* AD patients). The “former *vs*. latter” notation (*e.g*., Healthy vs. AD) is used to facilitate the presentation of SDP test results. Since the USs in healthy controls and AD patients represent distinct species assemblages in two separate treatments, their results are listed in separate tables. A second point worthy of mentioning is the SD (*q*=0), which measures species richness (number of species) within an assemblage. It is determined by the designation of species groups (*e.g*., US/ES) and is not weighted by species specificity. Therefore, SD (q = 0) comparisons are excluded from the SDP test discussion.

Key observations from the SD and SDP tests (Tables S19-S22) include:

(i) *US Species Assemblage:* 90% of pairwise comparisons between the five treatments (4 NNP disorders and healthy controls) showed significant differences in SD (Tables S19A and S19B).
(ii) *ES Species Assemblage:* 100% of pairwise comparisons showed significant differences in SD (Tables S20A and S20B).
(iii) *All Species with Significant Differences in Species Specificity:* 80% of pairwise comparisons showed significant differences in SD (Table S21).
(iv) *All Species (Without Considering Differences in SS):* 80% of pairwise comparisons showed significant differences in SD (Table S22).

In summary, the SDP tests revealed significant differences in SD for 80%-97% of pairwise comparisons between the five treatments (4 NNP disorders and healthy controls). These percentages are notably higher than those observed for microbial diversity differences between healthy and diseased treatments, which are approximately 1/3 for microbiome-associated diseases according to Ma *et al*. (2019).

### Neural Network and Machine Learning

As introduced in detail previously, we selected six AI-machine learning models—Neural Network, Random Forest, Support Vector Machine, Logistic Regression, Gradient Boosting, and K-Nearest Neighbor—and designed five schemes for organizing the training data and specifying the diagnosis schemes. These models were applied to differentiate gut microbiomes of healthy cohorts from those diagnosed with four neuropsychiatric and neurodegenerative (NNP) disorders: Alzheimer’s disease (AD), autism spectrum disorder (ASD), mood disorders (MD), and Parkinson’s disease (PD). In addition, we used 80% of the datasets for model-training and the remaining 20% for model-testing, repeating each modeling-testing process 100 times to ensure robustness of the results. Finally, we computed and reported the average precisions from the 100 repetitions.

Table S23-S26 (in MS-Excel) displays the training and test results for the five schemes. Table S27A-S27D summarizes the results from Table S23-S26. Key findings from these tables include the following: regarding,

i. Scheme I, which was designed to compare between the healthy cohort and each of the four disease cohorts individually (Healthy vs. AD, Healthy vs. ASD, Healthy vs. MD, and Healthy vs. PD). The mean precision (accuracy) ranged from 78.6 to 85.6 across the six algorithms. The Random Forest algorithm achieved the highest precision (85.6%). This scheme successfully distinguished each of the four NNP disorders from the healthy control (Fig 6A).
ii. Scheme II, which was designed to implement pairwise comparisons among the four disease cohorts (AD vs. ASD, AD vs. MD, AD vs. PD, ASD vs. MD, ASD vs. PD, and MD vs. PD). The average precision levels ranged from 81.5% to 96.8%. The Gradient Boosting algorithm demonstrated the highest performance (Fig 6B).
iii. Scheme III-A, which was designed to implement simultaneous classification of the four diseases (AD vs. ASD vs. MD vs. PD), exhibited average precision levels ranged from 0.286 (Neural network) to 0.936 (Gradient Boosting) (Fig 6C). Scheme III-B, which was designed to implement simultaneous classification of the four diseases (AD vs. ASD vs. MD vs. PD) plus the healthy control, exhibited average precision levels ranged from 0.054 (Neural network) to 0.647 (Gradient Boosting) (Fig 6D). The above-results are summarized from Table S23 (detailed MS-Excel tables) and Table S27A. These results are obtained from using *all* species (OTU) abundances information. A key takeaway from the results so far is that, using all species abundance data, the precision achievable with AI/machine learning for pairwise discrimination (Scheme I & II comparisons) can reach as high as 97% and generally exceeds 80%. Simultaneously discriminating among four diseases (Scheme III-A) can achieve precision levels as high as 94%, though it may drop to as low as 29%. However, a somewhat puzzling finding is that simultaneously discriminating among four diseases along with the healthy control (Scheme III-B) is more challenging than discriminating only the four diseases, with the maximal achievable precision dropping to approximately 65%. The remaining AI/machine learning results focus on testing the hypothesis that “less can be more” by restricting the training (and input) data to a small number of microbial species selected based on our prior SSD (species specificity and specificity diversity) analysis. Under Scheme V, the number of selected species is reduced to as few as approximately two dozen (specifically, 25).
iv. Scheme IV was designed to re-implement Schemes I and II using only the OTU abundance information of species that exhibited significant differences in species specificity—specifically, the unique and enriched species (US+ES). This approach aimed to test whether “less can be more” by leveraging US/ES abundance tables instead of the full OTU abundance tables for diagnosis. The detailed results from utilizing US/ES species only were reported in Table S24 and further summarized in Table S27B. Table S27B includes the precision values for this scheme and the delta (differences) in precision compared to Schemes I-II. For Scheme I, the delta values (precision of “less” scheme - precision of “more” scheme) across the six AI-ML models were [0.004, 0.000, 0.037, 0.007, −0.002, 0.011]. Notably, 5 out of 6 deltas were positive, with the only negative delta being negligible (−0.2%), indicating that “less” generally outperforms “more.” Similarly, for Scheme II, the delta vector was [0.007, 0.004, 0.037, −0.002, −0.001, 0.038], further supporting the superiority of the “less” approach. These results demonstrate that the precision improvement from using “less” can be as high as 4%, while the maximum precision loss is only 0.2%.
v. Scheme V ranks all species by species specificity (SS) in descending order and then selects the top 50 species (Scheme V-A) or top 25 species (Scheme V-B) with the highest SS to re-implement Schemes I, II, and III. Similar to Scheme IV, this scheme was designed to test whether “less can be more.” The detailed results from utilizing top 50 US/ES species only were reported in Table S25 and further summarized in Table S27C. Table S27C includes the precision values for this scheme and the delta (differences) in precision compared to Schemes I-III. Using the top 50 species, the delta values (precision of “less” scheme - precision of “more” scheme) for Scheme I across the six AI-ML models were [0.002, 0.000, 0.023, −0.009, −0.005, 0.012]. Here, 4 out of 6 deltas were positive, indicating that “less” is as good as “more.” For Scheme II, the delta vector was [0.009, 0.003, 0.038, −0.011, −0.002, 0.070], showing that “less” performs better, with precision improvements as high as 7%. For Scheme III-A, which involves simultaneously distinguishing all four NNP disorders (AD, ASD, MD, and PD) and healthy control, the delta vector was [-0.009, 0.005, 0.014, −0.004, −0.001, 0.04], 3 out of 6 deltas were positive, indicating that “less” is as good as “more”. Similarly, for Scheme III-B, which focuses on simultaneously distinguishing the four NNP disorders (without the added complexity of detecting the healthy control), the delta vector was [-0.037, 0.006, 0.021, −0.011, −0.002, 0.067]. The maximal gain in precision can reach up to 6.7%, while the maximal loss is 3.7%.

**Fig 6A.**
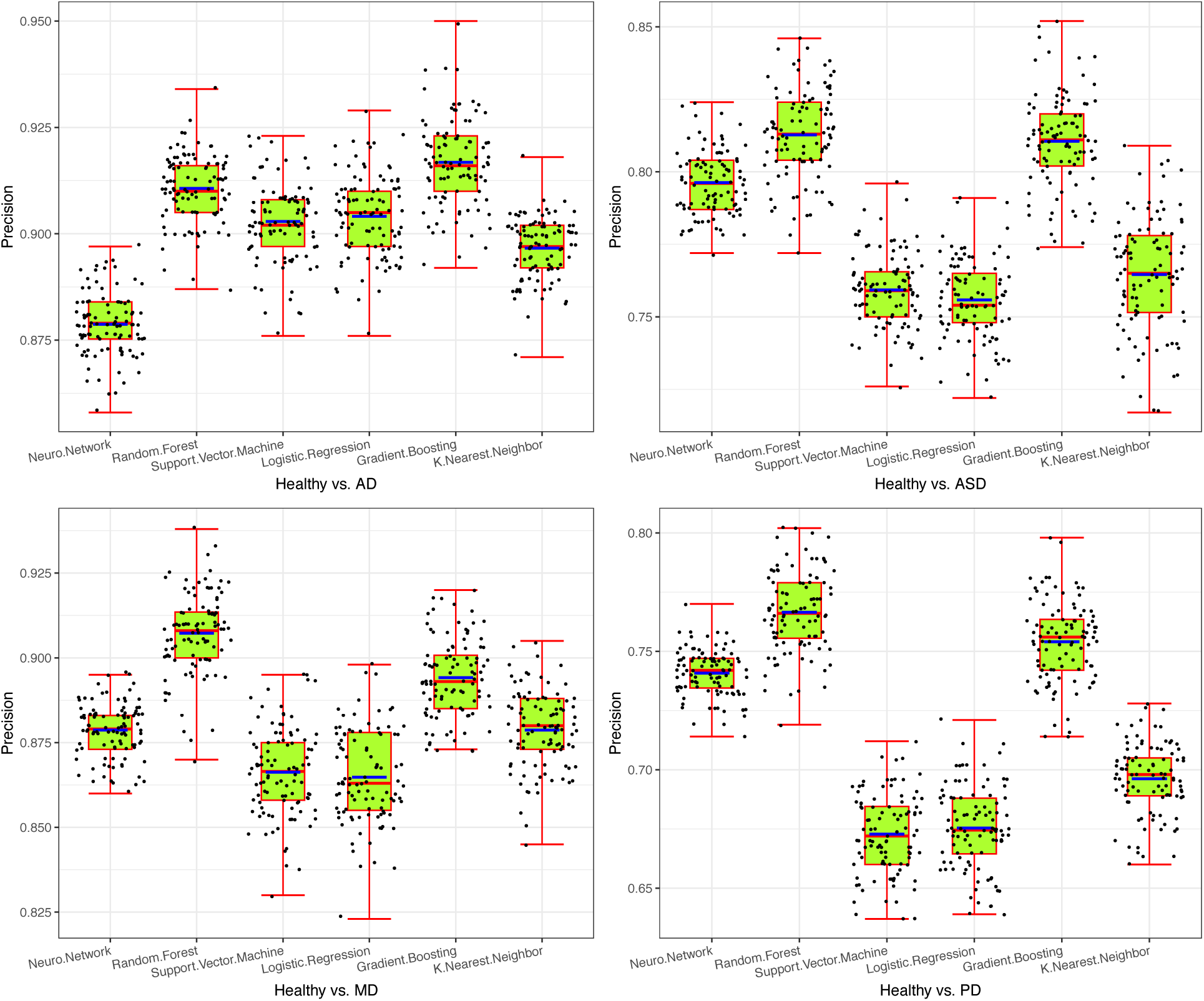
Performance evaluation (accuracy) of the six machine learning algorithms employed in Scheme-I (distinguishing each NNP disorder from the healthy control) using the top 25 species with the highest specificity values.

**Fig 6B.**
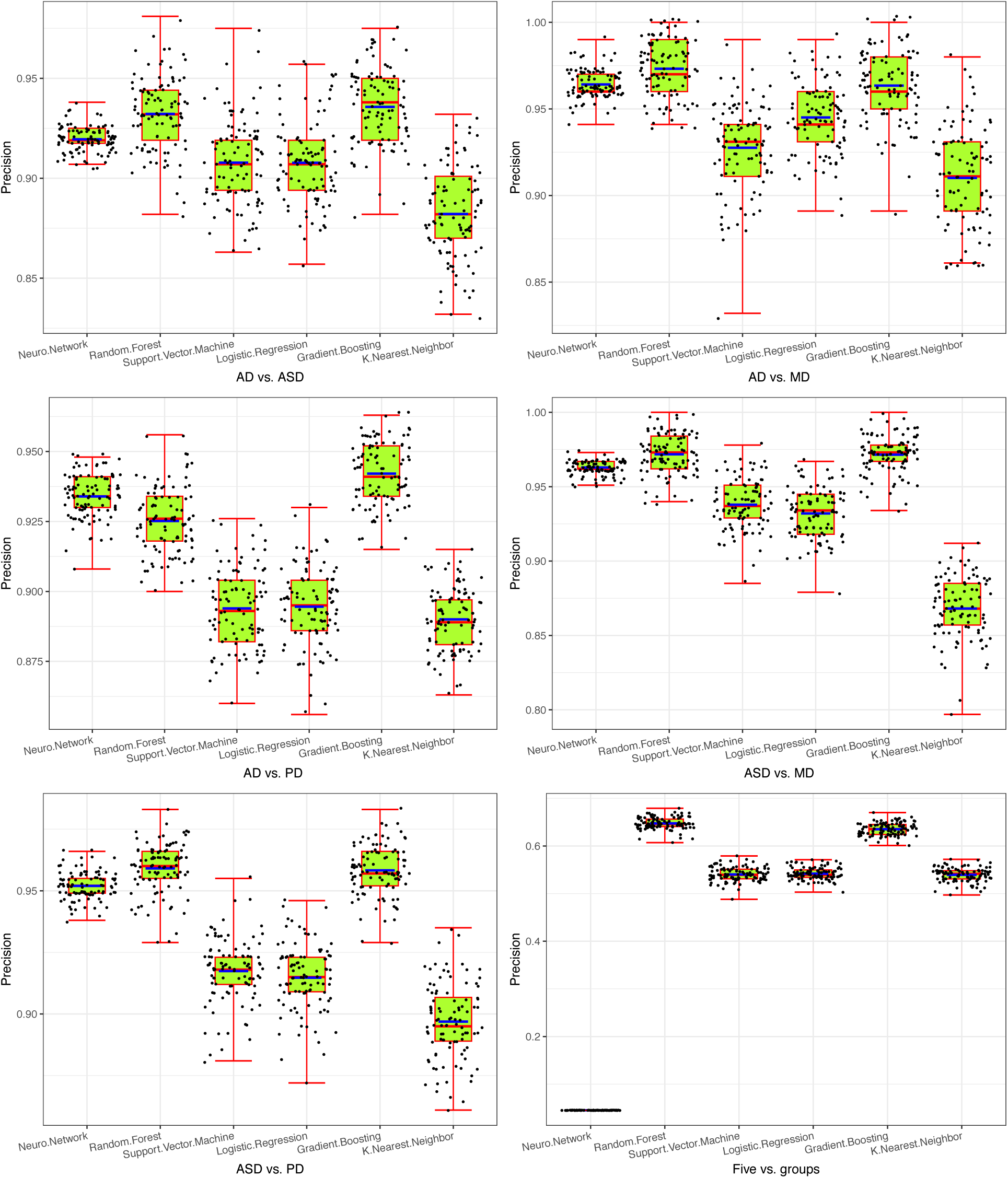
Performance evaluation (accuracy) of the six machine learning algorithms employed in Scheme-II (Pair-wisely discriminating four NNP disorder from each other) using the top 25 species with the highest specificity values.

**Fig 6C.**
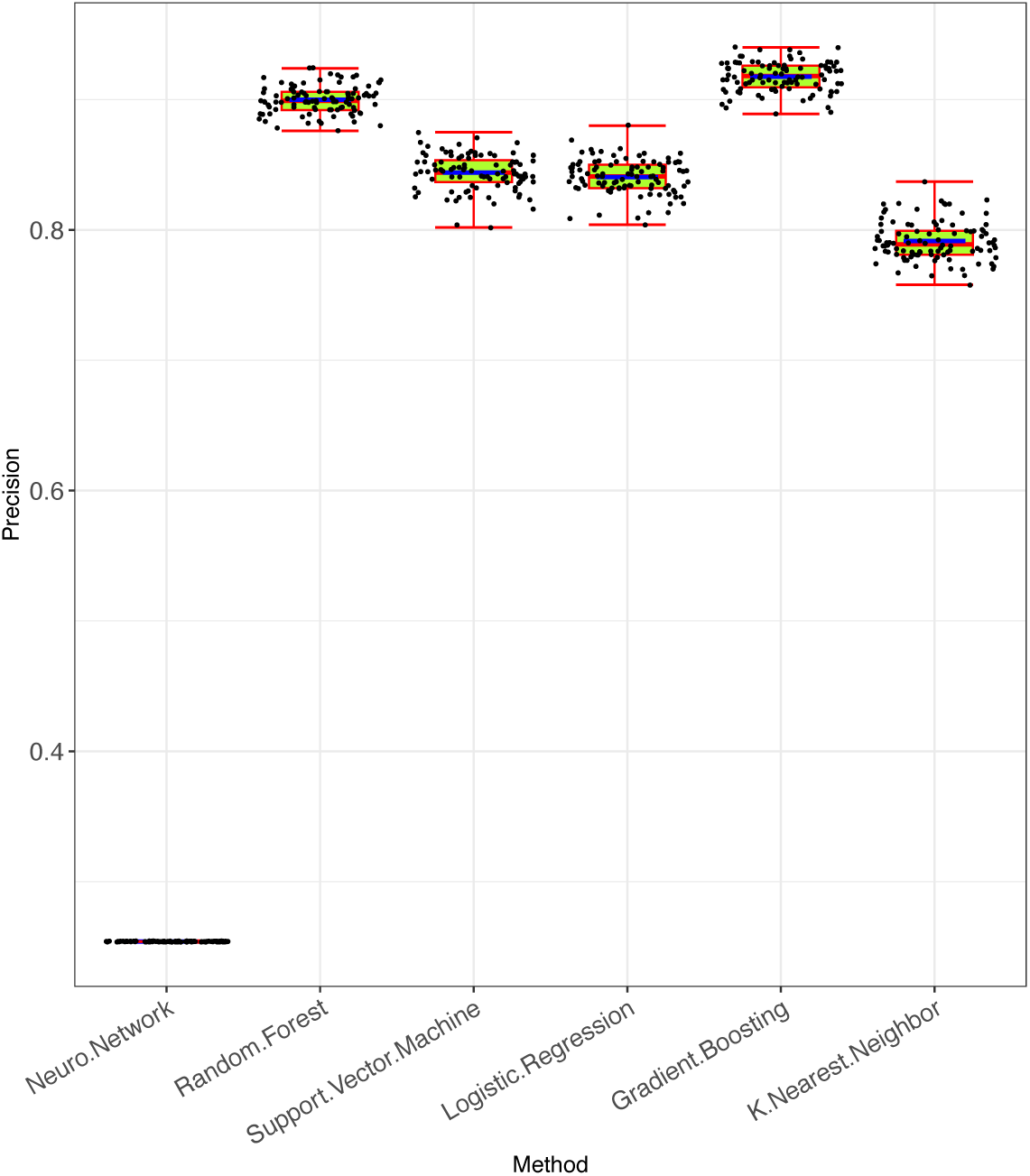
Performance evaluation (accuracy) of the six machine learning algorithms employed in Scheme-III-A (Discriminating four NNP disorders simultaneously) using the top 25 species with the highest specificity values.

**Fig 6D.**
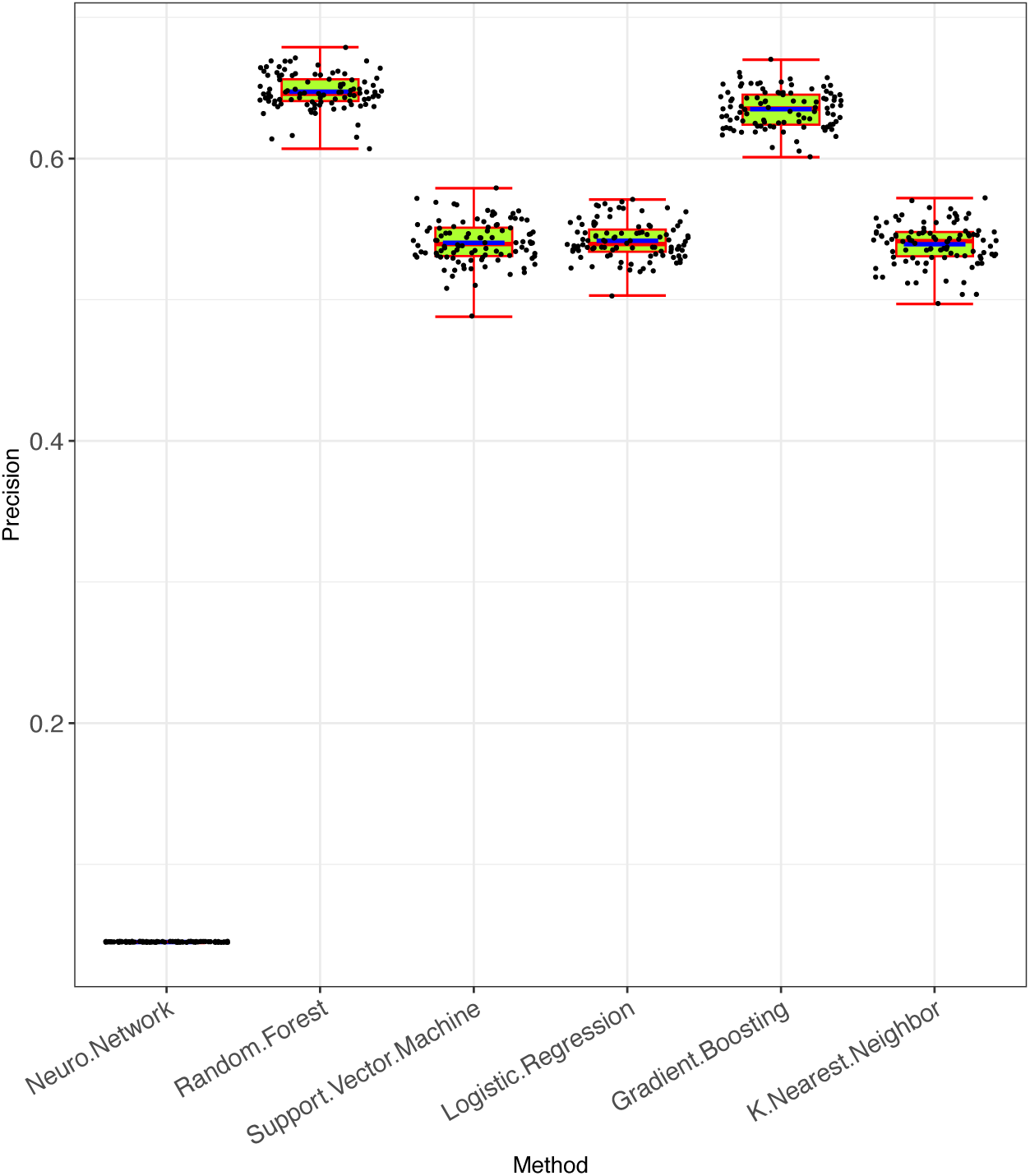
Performance evaluation (accuracy) of the six machine learning algorithms employed in Scheme-III-B (Discriminating healthy cohorts and four NNP disorders simultaneously) using the top 25 species with the highest specificity values.

To further investigate, we tested the top 25 species with Scheme V-B (Table S26 for the detailed results and Table S27D for the summary). For Scheme I, the delta vector was [-0.016, −0.007, 0.014, −0.019, −0.011, 0.012], indicating negligible precision loss, with a maximum loss of only 2%. For Scheme II, the delta vector was [0.001, −0.004, 0.025, −0.028, −0.010, 0.080], demonstrating that “less” can be as effective as “more,” with improvements reaching up to 8%. For Scheme III-A, the delta vector was [-0.009, −0.003, −0.019, −0.031, −0.012, 0.035]. Consistent with the results from the top 50 species, the performance with “less” (25 species) and “more” (all species) was remarkably similar—the maximum loss and gain were virtually identical (3.1% loss vs. 3.5% gain). For Scheme III-B, the delta vector was [-0.032, 0.000, −0.016, −0.04, −0.018, 0.082]. Similar to Scheme III-A, the precision remains virtually unchanged. The maximal gain in precision can reach 8.2%, while the maximal loss is 3.2%.

In summary, the results demonstrate that AI and machine learning algorithms are highly effective in differentiating between NNP disorders and healthy controls, as well as in simultaneously distinguishing among the four NNP disorders. Precision levels for these tasks generally exceed 80% and can reach as high as 97%. The only exception is the simultaneous discrimination of the four diseases along with the healthy control, which achieves a precision of approximately 65%. Even for the simultaneous discrimination of the four diseases alone (excluding healthy control), precision can reach up to 93%.

Even more compelling is the finding that “less can be more”—using smaller, carefully curated sets of OTU tables, with as few as 25 specialized species, can achieve comparable or even superior precision relative to using full OTU tables. To fully realize the benefits of this “less is more” approach, our SSD (species specificity and specificity diversity) framework plays an indispensable role in identifying the optimal subset of species. The lists of the top 25 species with the highest specificity (most “specific”) for each scheme, along with their species specificity values, are provided in Table S28. In fact, Fig. 6A-6D, which illustrate the precision of AI/machine learning models, were generated based on the results obtained using the top 25 species schemes.

Finally, the “less can be more” phenomenon underscores the critical role our SSD (species specificity and specificity diversity) analysis plays in effectively diagnosing NNP disorders using AI/machine learning approaches. Combined with the US/ES (unique/enriched species) lists identified through the SSD framework—which provide promising candidates for developing probiotics to treat NNP disorders—the SSD analysis offers valuable tools for both the diagnosis and treatment of these conditions.

## Conclusions and Discussion

### Conclusions

In recent years, numerous studies have explored alterations in the gut microbiome associated with neurological diseases. However, a systematic comparison of gut microbiome dynamics across different types of neurological disorders remains lacking. In this study, we present a comprehensive, comparative medical ecology analysis of four neurological disorders, encompassing neurodegenerative (Alzheimer’s disease [AD] and Parkinson’s disease [PD]), neurodevelopmental (autism spectrum disorder [ASD]), and psychiatric (depression [MD] and bipolar disorder [BD]) disorders, collectively referred to as NNP disorders. To achieve this, we employed a suite of four medical ecology approaches—diversity (incorporating alpha-, beta-, gamma-, and network diversity), heterogeneity (including dominance, abundance heterogeneity, and network heterogeneity), specificity (identifying disease-specific unique/enriched species), and AI/machine learning—selected to gain deep insights into dysbiosis at both microbiota and species levels in NNP disorders. Our findings include:

i. Comprehensive diversity analyses, from alpha-through beta-, gamma-, and network diversity, not only support existing consensus, such as altered diversity in NNP disorders, but also reveal new insights. For alpha-diversity, both disease type (DT) and health status (HS) significantly influence microbial diversity, though their interaction effects are insignificant. All NNP disorders increase alpha-diversity, with the specific order depending on species rarity (rare, common, or dominant). Beta-diversity analysis shows that all four NNP disorders follow the Anna Karenina Principle (AKP)—healthy individuals are more alike, while diseased ones are not. While diversity scaling across individuals is invariant across NNP disorders and the rarity-common-dominance spectrum, potential (gamma) diversity varies among disease types and the spectrum, particularly for dominant species. Network diversity (ND) of species relative abundance, link probability, and link abundance also varies with the rarity-common-dominance spectrum.
ii. Unlike diversity, a de facto standard analysis that ignores species interactions, heterogeneity— which considers both species abundance and interactions—has been understudied. Using TPLE (Taylor’s power law extension, emphasizing species abundance), heterogeneity scaling ranked as MD > ASD > PD - H > AD, while TPLoN (TPL of network, emphasizing species interactions) revealed an inverse order (H - PD > ASD > MD > AD), except for AD. Dominance analysis (emphasizing abundance asymmetricity) indicated the order of MD > PD > ASD - AD & D > H, similar to that of TPLE. These findings underscore the necessity of heterogeneity analysis— heterogeneity of species abundances and interactions can be rather different, while diversity ignores species interactions.
iii. The SSD (species specificity and specificity diversity) framework not only identified disease-specific unique/enriched species (US/ES), which are promising candidates for probiotic development, but also provided insights into the distinctiveness of each NNP disorder, supported by the previous diversity and heterogeneity analyses. Specifically, MD exhibited either negative correlations (with ASD, PD, and healthy controls) or insignificant specificity (with AD). PD and AD were positively correlated but not correlated with ASD, possibly due to patient age differences. These patterns align with intuitive perceptions of NNP disorders, though such perceptions lack microbiome-based evidence in existing literature. Furthermore, specificity diversity (SD) holistically differentiated between the NNP disorders (including healthy controls) in 80%-97% of pairwise comparisons, demonstrating higher discerning power than traditional diversity metrics.
iv. Machine learning algorithms, particularly random forest and neural networks, achieved 80%-97% precision in diagnosing NNP disorders.

Both the previous Specificity Diversity (SD) and machine learning algorithms achieved comparable levels of precision, further validating the feasibility of SSD (Species Specificity and Specificity Diversity) in diagnosing NNP disorders. Additionally, the previously mentioned US (Unique Species) and ES (Enriched Species), which were also identified through SSD analysis, provided valuable catalogues for developing potential probiotics to treat NNP disorders.

Even more encouraging is the finding that the number of microbial species required to distinguish between NNP disorders can be as few as approximately two dozen (25, to be exact). This remarkable specificity highlights the potential for developing streamlined diagnostic tools based on a relatively small set of microbial biomarkers. Such efficiency not only enhances the practicality of microbiome-based diagnostics but also underscores the feasibility of translating these findings into clinical applications. By focusing on a concise set of microbial signatures, future diagnostic approaches could become more accessible, cost-effective, and scalable, paving the way for earlier and more precise identification of NNP disorders.

These findings reveal shared and distinct microbiome features across neurodegenerative, neurodevelopmental, and psychiatric conditions, advancing our understanding of their etiological mechanisms. This knowledge can inform the development of microbiome-based diagnostic tools (e.g., microbial biomarkers) and targeted therapies (e.g., probiotics), paving the way for personalized interventions. By bridging gut microbiome ecology and neurological health, this study highlights the potential of medical ecology approaches to advance our understanding of complex neurological disorders and their microbiome-associated dynamics.

In the previous sections, we meticulously detailed the findings and insights derived from the four sets of medical ecology methods (frameworks)—diversity, heterogeneity, specificity, and machine learning. However, presenting a comprehensive panorama of the comparative analyses across the four NNP disorders and the healthy control remained challenging. To address this, Fig. 7 features nine pentagrams (five-pointed stars), with each of the four points representing an NNP disorder and the fifth point representing the healthy control. These pentagrams visually depict the differences/correlations (or lack thereof) among the five groups. The solid green lines indicate statistically significant differences, while the dashed grey lines represent the absence of such differences, as determined by rigorous statistical tests. In addition to the nine pentagrams in Fig. 7, two additional pentagrams are included as subgraphs in Fig. 3 and Fig. 5. Together, these 11 pentagrams encompass all the medical ecology metrics employed in this study. As such, Fig. 7 not only provides a panoramic overview of the study but also highlights minor details that may have been omitted in earlier sections.

**Fig. 7.**
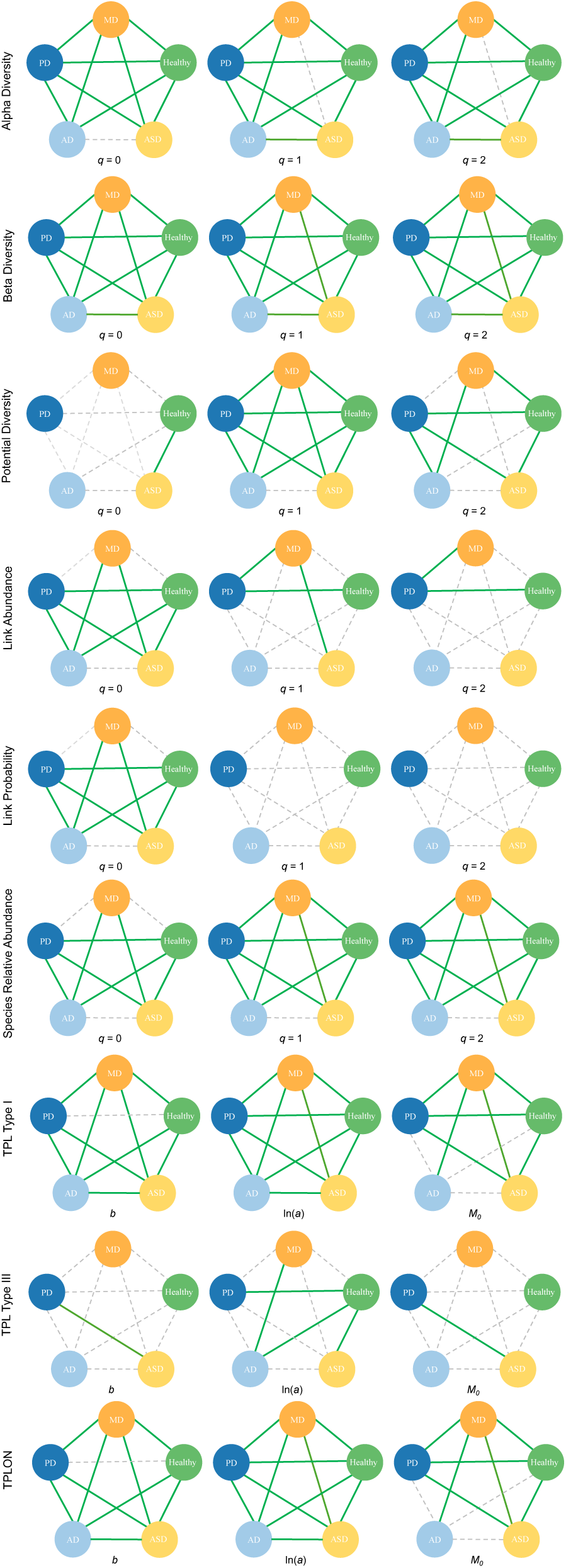
Summary Figures—Pentagrams of Pairwise Comparisons: The pentagrams summarize pairwise comparisons between healthy cohorts and the four NNP disorder cohorts based for nine indices of medical ecology analyses, respectively. Two additional pentagrams, presented in Fig. 3D (community dominance analysis) and Fig. 5D (specificity correlations), are omitted here for clarity. In the figures, the green line indicates a significant difference between the two groups, while the grey dotted line indicates no significant difference.

## Discussion

The gut microbiome has been increasingly recognized as a critical factor in the pathogenesis of neurodegenerative, neurodevelopmental, and psychiatric disorders (collectively referred to as NNP disorders). In this study, we utilized a comprehensive medical ecology framework to analyze the gut microbiome in individuals with Alzheimer’s disease (AD), Parkinson’s disease (PD), autism spectrum disorder (ASD), and mood disorders (MDs). By integrating diversity, heterogeneity, specificity, and machine learning approaches, we identified both shared and distinct microbial features across these disorders, offering new insights into their underlying mechanisms and potential therapeutic targets. Below, we synthesize our findings with the broader literature, emphasizing key themes and their implications for future research and treatment strategies.

### Diversity and Dysbiosis in NNP Disorders

Our findings align with existing literature demonstrating altered microbial diversity in NNP disorders. For instance, Liu et al. (2020) reported reduced levels of anti-inflammatory, butyrate-producing bacteria (e.g., *Faecalibacterium*) in individuals with major depressive disorder (MDD), consistent with our observation of increased alpha-diversity in MD. Similarly, Zhou et al. (2020) found reduced *Firmicutes* and increased *Enterobacteriaceae* in postpartum depression, mirroring our findings of disease-specific shifts in microbial composition. These changes in diversity are not merely correlative but may reflect underlying pathophysiological mechanisms, such as inflammation and impaired gut-brain axis communication.

In neurodegenerative disorders, Zhuang et al. (2018) and Ling et al. (2021) reported distinct microbial profiles in AD patients, including reduced *Faecalibacterium* and increased *Bacteroides*, which align with our specificity analysis identifying disease-enriched species. Similarly, Aho et al. (2019) and Weis et al. (2019) highlighted temporal stability and medication effects on gut microbiota in PD, underscoring the importance of longitudinal studies in understanding microbial dynamics.

### Heterogeneity and Species Interactions

While diversity metrics provide a broad overview of microbial communities, heterogeneity analyses offer deeper insights into species interactions and abundance patterns. Our study revealed significant differences in heterogeneity scaling across NNP disorders, with MD exhibiting the highest TPLE (Taylor’s power law extension) values, emphasizing the role of species abundance variability. This finding resonates with Hu et al. (2019), who observed lower levels of butyrate-producing bacteria in bipolar depression, suggesting that microbial heterogeneity may reflect functional disruptions in metabolic pathways.

The inverse order observed in TPLoN (Taylor’s power law of networks) further highlights the importance of species interactions. For example, PD and healthy controls showed similar network heterogeneity, consistent with Aho et al. (2019), who reported stable microbial communities in PD over time. These findings suggest that while diversity metrics capture overall community structure, heterogeneity analyses provide a more nuanced understanding of microbial dynamics.

### Disease-Specific Microbial Signatures

Our SSD (species specificity and specificity diversity) framework identified unique and enriched species (US/ES) across NNP disorders, offering potential biomarkers for diagnosis and therapeutic targets. For instance, MD exhibited negative correlations with ASD, PD, and healthy controls, consistent with Liu et al. (2020), who reported reduced *Faecalibacterium* in MDD. Similarly, PD and AD showed positive correlations, aligning with Zhuang et al. (2018) and Ling et al. (2021), who identified shared microbial alterations in these disorders.

In neurodevelopmental disorders, Zurita et al. (2020) and Pulikkan et al. (2018) reported increased *Bacteroides* and *Akkermansia* in ASD, consistent with our findings of disease-specific enrichment. These microbial signatures may reflect underlying genetic and environmental factors, as highlighted by Liu et al. (2021), who identified associations between host genetic variations and gut microbiota in ASD.

### Machine Learning and Diagnostic Potential

Machine learning algorithms achieved high precision in diagnosing NNP disorders, underscoring the potential of microbiome-based biomarkers. For example, Hu et al. (2019) identified microbial markers with high diagnostic accuracy for bipolar disorder, while Wallen (2021) compared differential abundance testing methods, emphasizing the importance of robust statistical frameworks. These findings suggest that integrating microbial data with clinical parameters could enhance diagnostic precision and facilitate personalized interventions.

### Implications for Therapeutic Interventions

The identification of disease-specific microbial signatures and functional pathways opens new avenues for therapeutic interventions. Probiotics targeting *Faecalibacterium* and other butyrate-producing bacteria could mitigate inflammation and restore gut-brain axis function in MD and AD. Similarly, dietary interventions modulating *Bacteroides* and *Akkermansia* levels may benefit individuals with ASD, as suggested by Zurita et al. (2020) and Pulikkan et al. (2018).

Moreover, our findings highlight the potential of microbial biomarkers for monitoring disease progression and treatment response. For instance, Aho et al. (2019) observed microbial changes following quetiapine treatment in PD, suggesting that gut microbiota could serve as a dynamic indicator of therapeutic efficacy.

### Limitations and Future Directions

While this study provides a comprehensive analysis of gut microbiome dynamics in NNP disorders, several limitations warrant consideration. First, the cross-sectional design precludes causal inferences, necessitating longitudinal studies to elucidate temporal relationships. Second, the heterogeneity of patient populations, including age, medication use, and comorbidities, may confound microbial analyses. Future studies should stratify patients based on these factors to identify more precise microbial signatures.

Additionally, the integration of multi-omics data (e.g., metagenomics, metabolomics) could provide deeper insights into functional pathways and host-microbe interactions. For example, Liu et al. (2021) and Ding et al. (2020) highlighted the role of microbial metabolites in ASD, suggesting that combining microbial and metabolic profiling could enhance our understanding of disease mechanisms.

In conclusions, this study advances our understanding of the gut microbiome’s role in NNP disorders by identifying shared and distinct microbial features across AD, PD, ASD, and MD. By integrating diversity, heterogeneity, specificity, and machine learning approaches, we provide a robust framework for analyzing microbial dynamics and their clinical implications. These findings underscore the potential of microbiome-based diagnostics and therapeutics, paving the way for personalized interventions that target the gut-brain axis. Future research should focus on longitudinal studies, multi-omics integration, and clinical trials to validate these findings and translate them into actionable strategies for improving neurological health.

## Data Availability

All data reanalyzed in this study are publicly available in the NCBI database. Detailed access information is provided in the manuscript.

## Standard Declarations

### Data and Code Accessibility Statement

All data reanalyzed in this study are publicly available, with access details provided in Table S1. The computational codes are also publicly accessible and can be found in the respective method papers cited in the Materials and Methods section.

### Author Contributions

ZSM conceived and designed the study, developed the methodology, interpreted the results, wrote and revised the manuscript. QYT and LLW were responsible for the data curation and data analysis, contributed to the interpretation of the results, and prepared the tables and figures.

### Funding and Acknowledgements

The study received funding from the following sources: National Natural Science Foundation (NSFC #31970116, #72274192); 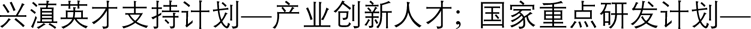,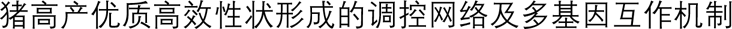; Bullard Fellowship from Harvard University, USA.

